# Reduced effectiveness of repeat influenza vaccination: distinguishing among within-season waning, recent clinical infection, and subclinical infection

**DOI:** 10.1101/2023.03.12.23287173

**Authors:** Qifang Bi, Barbra A. Dickerman, Huong Q. Nguyen, Emily T. Martin, Manjusha Gaglani, Karen J. Wernli, G.K. Balasubramani, Brendan Flannery, Marc Lipsitch, Sarah Cobey, the US Flu Vaccine Effectiveness Network Investigators

**Affiliations:** University of Chicago, Chicago, Illinois, USA; Harvard T.H. Chan School of Public Health, Boston, Massachusetts, USA; Center for Clinical Epidemiology & Population Health, Marshfield Clinic Research Institute, Marshfield, Wisconsin, USA; University of Michigan School of Public Health, Ann Arbor, Michigan, USA; Baylor Scott & White Health, Temple, Texas, USA; Texas A&M University College of Medicine, Temple, Texas, USA; Kaiser Permanente Bernard J. Tyson School of Medicine, Seattle, Washington, USA; University of Pittsburgh School of Public Health, Pittsburgh, Pennsylvania, USA; National Center for Immunization and Respiratory Diseases, Centers for Disease Control and Prevention, Atlanta, GA, USA

**Keywords:** influenza, vaccine, waning vaccine protection, infection history, infection block hypothesis, immunogenicity, test negative design

## Abstract

Studies have reported that prior-season influenza vaccination is associated with higher risk of clinical influenza infection among vaccinees. This effect might arise from incomplete consideration of within-season waning and recent infection. Using data from the US Flu Vaccine Effectiveness (VE) Network (2011-2012 to 2018-2019 seasons), we found that repeat vaccinees were vaccinated earlier in a season by one week. After accounting for waning VE, repeat vaccinees were still more likely to test positive for A(H3N2) (OR=1.11, 95%CI:1.02-1.21) but not for influenza B or A(H1N1). We found that clinical infection influenced individuals’ decision to vaccinate in the following season while protecting against clinical infection of the same (sub)type. However, adjusting for recent clinical infections did not strongly influence the estimated effect of prior-season vaccination. In contrast, we found that adjusting for subclinical infection could theoretically attenuate this effect. Additional investigation is needed to determine the impact of subclinical infections on VE.

Summary of main points: Two potential factors, timing of vaccination and clinical infection history, cannot fully explain the increased influenza infection risk in repeat vaccinees compared with non-repeat vaccinees. Subclinical infection in the previous season may explain the effect.

## 2 Introduction

The World Health Organization recommends annual influenza vaccination of persons at high risk, with some countries recommending universal vaccination[1,2]. A controlled study in the 1970s first raised questions about repeated annual influenza vaccination, reporting that prior vaccination indirectly increased the risk of infection in the current season[3,4]. It was not until a test-negative study in Canada[5], a vaccine trial in Hong Kong[6] and a household-based study in the United States[7] found differences in vaccine effectiveness (VE) and immunogenicity among repeat vaccinees and non-repeat vaccinees in the 2009/10 and 2010/11 seasons that the phenomenon was investigated routinely[8–14]. Since then, increased infection risk against A(H3N2) in repeat vaccinees was observed in multiple seasons and countries[7,11,12,13,15,16]. Increased risk is less often reported for the less prevalent A(H1N1) and type B[8,9].

Test-negative studies conducted in healthcare settings have become the standard way to evaluate vaccine protection. A test-negative design estimates VE by comparing vaccination coverage in persons with a medically attended acute respiratory illness who test positive for influenza with those who test negative[17]. Several factors that may bias estimates of repeat vaccination effects in test-negative design have not been considered.

Vaccine-induced protection against influenza virus infection wanes within a season[18–22]. Consequently, the vaccine protection estimated among otherwise similar vaccinees may differ if the timing of vaccination is not considered. If repeat vaccinees tend to vaccinate substantially earlier in a season, waning protection could make the risk of infection among repeat vaccinees appear higher than in non-repeat vaccinees. The rapidly changing risk of influenza incidence in a season may amplify the difference[23].

The infection block hypothesis[4,24–26] suggests that prior vaccinations can block opportunities to experience immunogenic influenza virus infections, which can lead to more cross-reactive and durable immune responses than vaccination[27], especially when circulating viruses differ from vaccine strains[28,29]. If true, the infection-block hypothesis could explain increased risk of infection among repeat vaccinees compared to non-repeat vaccinees: Prior-season vaccination could protect repeat vaccinees against prior-season infection, leaving them less immune protection at the start of an influenza season and hence a higher incidence of clinical infection in that season than non-repeat vaccinees. The difference in risk between the two groups can be further amplified if recent infection improves vaccine immunogenicity and vaccine-induced protection[28], as has also been recently observed for SARS-CoV-2[30].

In epidemiologic terms[31], under the infection block hypothesis, infection in the previous season is a mediator between vaccination in the previous season and a clinical infection outcome in the current season. When we estimate the effect of repeated vaccination, infection in the previous season, acting as a mediator, does not inherently introduce bias. However, if infection in the previous season influences the decision to vaccinate in the current season as well as the probability of clinical infection in the current season, then it is also a confounder that can bias the estimated effect of repeated vaccination on clinical infection. Because infection in the previous season may be both a mediator and a confounder, appropriately adjusting for it requires an approach that can handle this treatment-confounder feedback, such as inverse-probability weighting[32].

In this study, we first assessed the effect of repeated vaccination after accounting for intra-season waning of vaccine protection (results in Section 4.1). We then assessed whether *clinical* infection in the prior season, a potential confounder, may have biased our estimate of the effect of repeated vaccination (Section 4.2). Finally, we theoretically assessed the plausibility of the infection block hypothesis and enhanced VE from recent *subclinical* infections as explanations for the repeat vaccination effect (Section 4.3).

## 3 Methods

### 3.1 Study setting and population

During the study period, the US Flu VE Network consisted of five study sites in Wisconsin, Michigan, Washington, Pennsylvania, and Texas[7,10,33,34,35] (Supplemental Section 1). The study used a test-negative design, estimating the odds of influenza infection in individuals who were vaccinated vs unvaccinated. During each enrollment season, outpatients 6 months of age and older were eligible for recruitment if they presented with acute respiratory illness with symptom onset within the last 7 or 10 days, depending on the Flu VE site. Each eligible patient completed an enrollment interview that included questions on status of influenza vaccination in the study enrollment season (the current season), influenza vaccination in the immediately preceding season (the previous season), demographic information, and underlying health conditions. Participants were tested for influenza by real-time reverse transcription polymerase chain reaction (rRT-PCR) assay. Influenza-positive samples were first typed and then A-sub- or B-lineage-typed. For simplicity throughout, we refer to individuals with medically attended PCR-confirmed symptomatic influenza virus infection as having “clinical infection”. Influenza vaccination status was confirmed by reviewing immunization records and state registries.

We analyzed data collected over 8 seasons (from the 2011-2012 through the 2018-2019 seasons) from all five sites. We excluded individuals who were vaccinated within 14 days of illness onset, for consistency with prior analyses. We excluded individuals who received more than one dose each season before symptom onset and were under 1 year of age at enrollment.

To study the impact of clinical infection history, we additionally obtained enrollment history and rRT-PCR testing history from the Marshfield Clinic (MCHS), the US Flu VE Network site in Wisconsin. The study design and the definition of clinical infection were consistent over time. As the primary outpatient and inpatient care provider in its catchment area, MCHS could collect data on enrollment and testing history that are not available from other sites [36]. In particular, participant data are linked across seasons. We analyzed data from the MCHS over 12 seasons (the 2007-2008 through the 2018-2019 seasons). The analyses using exclusively MCHS data are described in the subsection ‘Adjustment for clinical infection history’ of Section 3.2, and the results are shown in Section 4.2.

### 3.2 Statistical analyses

#### Accounting for within-season waning of vaccine protection

Using data from the five sites in the US Flu VE Network, we first determined whether the timing of vaccination differed between repeat and non-repeat vaccinees by fitting a linear regression model.

Using logistic regression models, we then estimated the relative odds of clinical infection among repeat vaccinees with reference to non-repeat vaccinees after adjusting for time of vaccination in the current season (to account for the waning of vaccine protection; Supplemental Section 3). The study outcome is (sub)type-specific PCR-confirmed clinical infection. Independent variables are an indicator for having been vaccinated 2-9, 10-13, 14-17, 18-21, or over 21 weeks before symptom onset in the current season regardless of prior-season vaccination status (categorization consistent with Ray *et al.*[18]), a dichotomous indicator for having been vaccinated only in the prior season, a dichotomous indicator for having been vaccinated in both the current and the prior season, age group, sex, comorbidity, influenza season, study site, and calendar month of symptom onset.

#### Adjustment for clinical infection history

Because MCHS was the only site that had linked participants’ previous study enrollment and infection history, only data from MCHS could be used to assess the impact of clinical infection history.

To determine how a clinical infection in the current season is associated with clinical infection with the same and other (sub)types in prior seasons, we assessed the odds ratio of clinical infection in the current season among individuals with no prior clinical infections or clinical infections 3-5 seasons or ≥6 seasons ago with reference to those whose last detected clinical infection was 1-2 seasons before the current season (Supplemental Section 4).

We then assessed whether clinical infections in the previous season influenced the decision to vaccinate in the current season using logistic regression models. The dependent variable was vaccination in the current season. The model was stratified by previous-season vaccination status, and additionally adjusted for age group, sex, comorbidity, and an indicator of vaccination frequency(Supplemental Section 4).

Next, we estimated the effect of repeated vaccination after adjusting for the clinical infection status of any (sub)type in the previous season (Supplemental Section 4). To handle the treatment-confounder feedback, we used inverse-probability weighting to account for clinical infection status of any (sub)type in the previous season, using regression to adjust for baseline covariates. Weights were calculated as the inverse of each individual’s probability of being vaccinated in each season given their previous vaccination status, infection status in the prior season, and baseline covariates (i.e., sex, age group, comorbidities, influenza season). These weights were then “stabilized” using the probabilities of being vaccinated given vaccination history, and the above-mentioned baseline covariates (excluding infection status).

#### Impact of subclinical infection

To understand how subclinical infection history, i.e., infections not detected by the US Flu VE Network, may impact the estimated effect of repeated vaccination, we evaluated the proportion of repeat and non-repeat vaccinees who would have had to have been subclinically infected in the previous season to reproduce the estimated effect of repeated vaccination, assuming that the subclinical infection-block hypothesis was the only explanation for the observed elevated risk. We demonstrate that the results are consistent with the hypothesis of enhanced vaccine immunogenicity post-infection.

To achieve this objective, we built a theoretical model and created a pseudo-population of repeat and non-repeat vaccinees with various infection statuses in the previous seasons (Supplemental Section 5). We derived the relationship between rates of subclinical infections in repeat- and non-repeat vaccinees given various degrees of effectiveness of subclinical infection against future clinical infection (30%, 50%, and 70% reductions in clinical infection risk). We varied assumptions about the protection conferred by clinical and subclinical infection in the prior season against future infection. Based on estimates from prior studies[37,38], we varied clinical attack rates in vaccinated and unvaccinated individuals, assuming either low (1% and 2% for vaccinated and unvaccinated individuals, respectively) or high clinical incidence (3% and 6% respectively).

The study obtained the institutional review board approval at participating institutions and the Centers for Disease Control and Prevention.

## 4 Results

Between the 2011-2012 and 2018-2019 seasons, individuals enrolled in the US Flu VE Network contributed 61,943 visits, of which 55,728 (90.0%) met the inclusion criteria of our analyses. Of those, 50.2% (27,986/55,728) of visits were by individuals who had received one dose of the current seasonal influenza vaccine _≥_14 days prior to illness onset date (SFig 1.1). Among those vaccinated _≥_14 days prior to illness onset, 73.7% (20,630/27,986) of visits were by individuals who were vaccinated at least once in the previous season, and whom we refer to as repeat vaccinees (STable 1.1).

**Figure 1:**
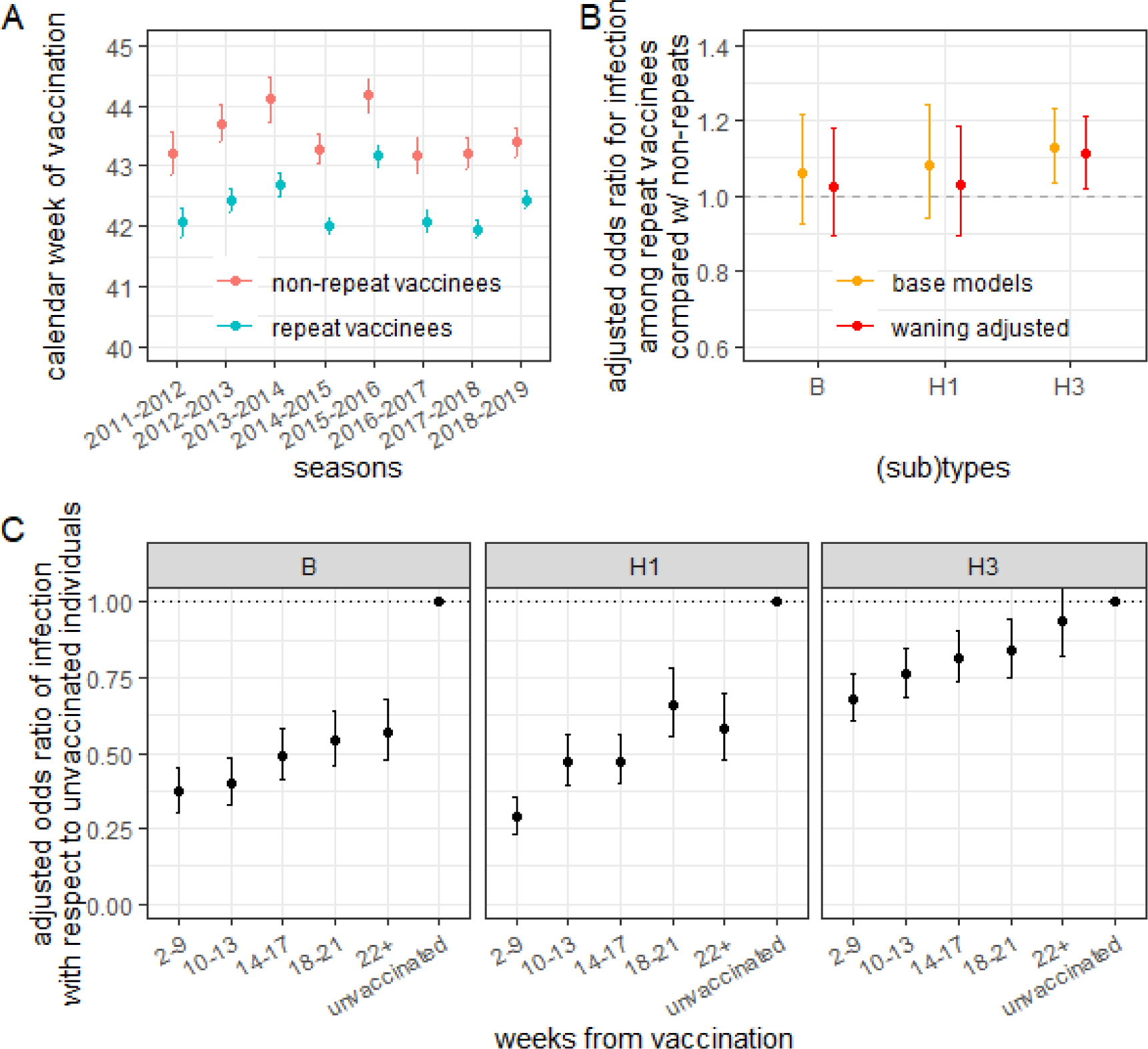
That repeat vaccinees vaccinate earlier in a season, which increases their susceptibility to infection due to waning vaccine protection, does not explain their higher odds of infection compared to non-repeat vaccinees. A) Average calendar week of vaccination among repeat and non-repeat vaccinees over the study enrollment seasons. Repeat vaccinees consistently get vaccinated earlier than non-repeat vaccinees. B) Adjusted odds ratio for clinical infection among individuals vaccinated this season stratified based on whether the individuals were also vaccinated in the prior season (repeat vaccinees) or not (non-repeat vaccinees) before (yellow) and after (red) adjusting for the timing of vaccination within a season. Site- and season-specific data are shown in SFig 3.3). C) Adjusted odds ratio of clinical infection comparing individuals vaccinated 2-9, 10-13, 14-17, 18-21, and 22+ weeks in the current season (but not in the prevoius season) before testing positive with respect to those not vaccinated in either season. Site- and season-specific data are shown in SFig 3.1 and age specific data are shown in SFig 3.5. See Supplemental Section 3 for detailed definitions of the quantities reported here.

### 4.1 Impact of waning vaccine protection

On average, repeat vaccinees of similar age, sex, and comorbidities were vaccinated 1.1 (95%CI:1.0-1.2) weeks earlier than non-repeat vaccinees (Figure 1A). Adjusting for the timing of vaccination in the current season did not notably change the marked repeat vaccination effect for A/H3N2 and had little to no effect for A/H1N1pdm09 and type B (Figure 1B; SFig 3.3 shows variation in estimates by season and site; SFig 3.5 shows results did not vary significantly by age group).

In models accounting for the timing of vaccination and previous season vaccination, we observed that odds of infection against all three (sub)types increased with time since current season vaccination (Figure 1C). Compared with individuals not vaccinated in either season (who had the highest risk of testing positive), current-season vaccinees who vaccinated 2-9 weeks before testing had lower OR (0.29 [95%CI:0.23-0.35]) for A/H1N1pdm09-associated illness than those vaccinated 18-21 weeks before testing (OR=0.66; 95%CI:0.56-0.78). In the 2014-2015 season, when there was a mismatch between the A/H3N2 component and the circulating strains, the odds of infection decreased with time from vaccination in Wisconsin (SFig 3.1).

### 4.2 Impact of clinical infection history

Prior clinical infections of the homologous (sub)type protected against clinical infections of type B or A/H3N2, with more recent infections conferring stronger protection (Figure 2A); those infected with type B more than 6 seasons ago had 3.60 (95%CI:1.08-11.9) times the odds of testing positive for type B in the current season than those who were clinically infected in the previous 1-2 seasons (Figure 2A). A similar trend emerged for clinical infections against A/H3N2 (OR=32.4, 95%CI:4.4-242, Figure 2A). We did not find clinical infections of a heterologous (sub)type to be protective (SFig 4.1). Due to the limited number of A/H1N1pdm09 infections during our enrollment period, we could not assess the impact of homologous infection with A/H1N1pdm09.

**Figure 2:**
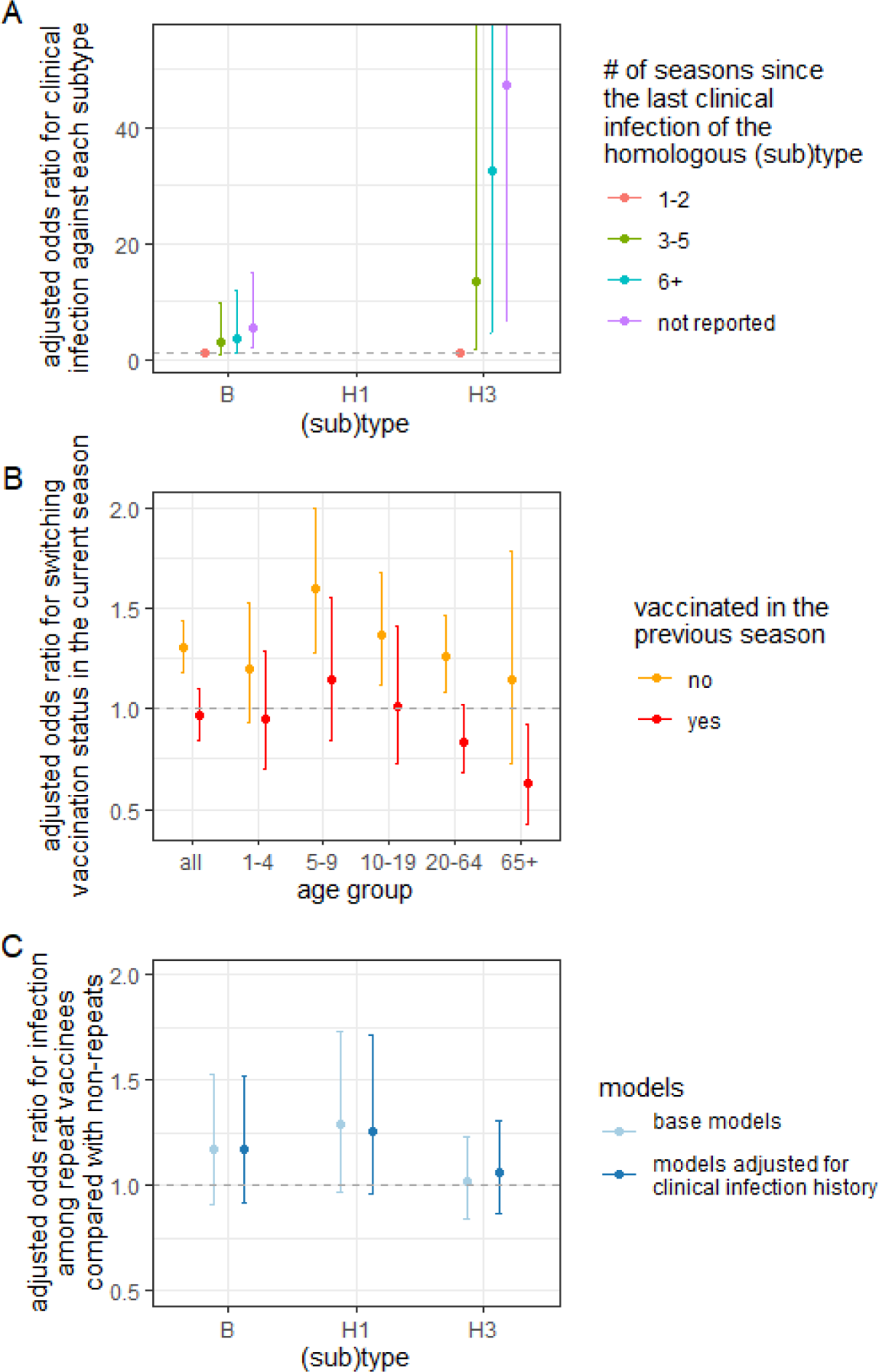
Recent clinical infections, which induce non-vaccinees to vaccinate the next season and which can protect against clinical reinfection for years, cannot explain the effect of repeated vaccination. A) Association between recent clinical infections and odds of current-influenza-season clinical infection. More distant clinical infections of the homologous subtype are associated with a higher odds of current-season clinical infection. B) Tendency to switch vaccination status in the current influenza season after clinical infection in the previous season. Compared with individuals without confirmed infections, unvaccinated individuals who were clinically infected in the previous season were more likely to vaccinate in the current season. C) Estimated effect of repeat vaccination after adjusting for recent clinical infections. Adjusted odds ratio for clinical infection comparing repeat vaccinees with non-repeat vaccinees before (light blue) and after adjusting for clinical infection status in the previous season (dark blue) using inverse-probability weighting. Results stratified by age group are shown in SFig 4.4. They suggest a marginally significantly higher adjusted odds of infection in repeat vaccinees >19 years old for H1N1. Adjustment did not significantly impact the estimates. In all panels, error bars indicate 95% confidence intervals.

We found that having a confirmed influenza virus infection in the previous season appeared to influence the decision to vaccinate in the current season. Individuals unvaccinated in the previous season were more likely to vaccinate in the current season (OR=1.30, 95%CI:1.18-1.44) if they were clinically infected in the previous season than if they were not infected. However, individuals who became infected after being vaccinated in the previous season were as likely to be unvaccinated in the current season as those not infected (OR=0.96, 95%CI:0.85-1.10), with the exception of the oldest age group, which tended to vaccinate again (Figure 2B).

Adjusting for confounding by clinical infection in the previous season had little influence on the estimated effect of repeated vaccination (Figure 2C). After adjustment, repeat vaccinees enrolled during the 2008-2009 season and between the 2010-2011 and the 2018-2019 seasons had 1.29 (95%CI:0.96-1.71) times the odds of testing positive for A/H1N1dpm09 than those who were only vaccinated in the current season. Accounting for clinical infection history did not significantly change the estimated effect of repeated vaccination against A/H3N2 (from 1.02, 95%CI:0.84-1.23 to 1.06, 95%CI:0.87-1.31 post adjustment) or type B (from 1.18, 95%CI:0.91-1.53 to 1.17, 95%CI:0.92-1.52 post adjustment). Excluding the 194 individuals who presented with acute respiratory illness but refused enrollment in the previous season did not significantly change the results (SFig 4.2). Not adjusting for waning vaccine protection in the weighted outcome model yielded similar results (SFig 4.3, Supplemental Section 4).

### 4.3 Impact of clinical and subclinical infection history

#### 4.3.1 Infection block hypothesis

In the previous section we estimated that repeat vaccinees had a 10% increase (OR∼1.1) in the odds of current-season infection, an effect that could be partially mediated by clinical infection in the prior season, a version of the infection block hypothesis. In this section, we use a theoretical model to explore the degree to which subclinical infection – which would not be observed in any of the data sets we consider – could fully explain the observed repeat vaccination effect.

To produce the estimated effect of repeated vaccination (i.e., OR for clinical infection comparing repeat with non-repeat vaccinees against A/H3N2 or type B of 1.1) in the US Flu VE Network, non-repeat vaccinees would have to be subclinically infected in the prior season at a substantially higher rate than repeat vaccinees (Figure 3; SFig 5.1). For example, if subclinical infection reduces the probability of next-season clinical infection by 70% (dark green curve in Figure 3A), ∼5% of repeat vaccinees and ∼15% of non-repeat vaccinees would have to have been subclinically infected in the prior season to observe the estimated effect.

**Figure 3:**
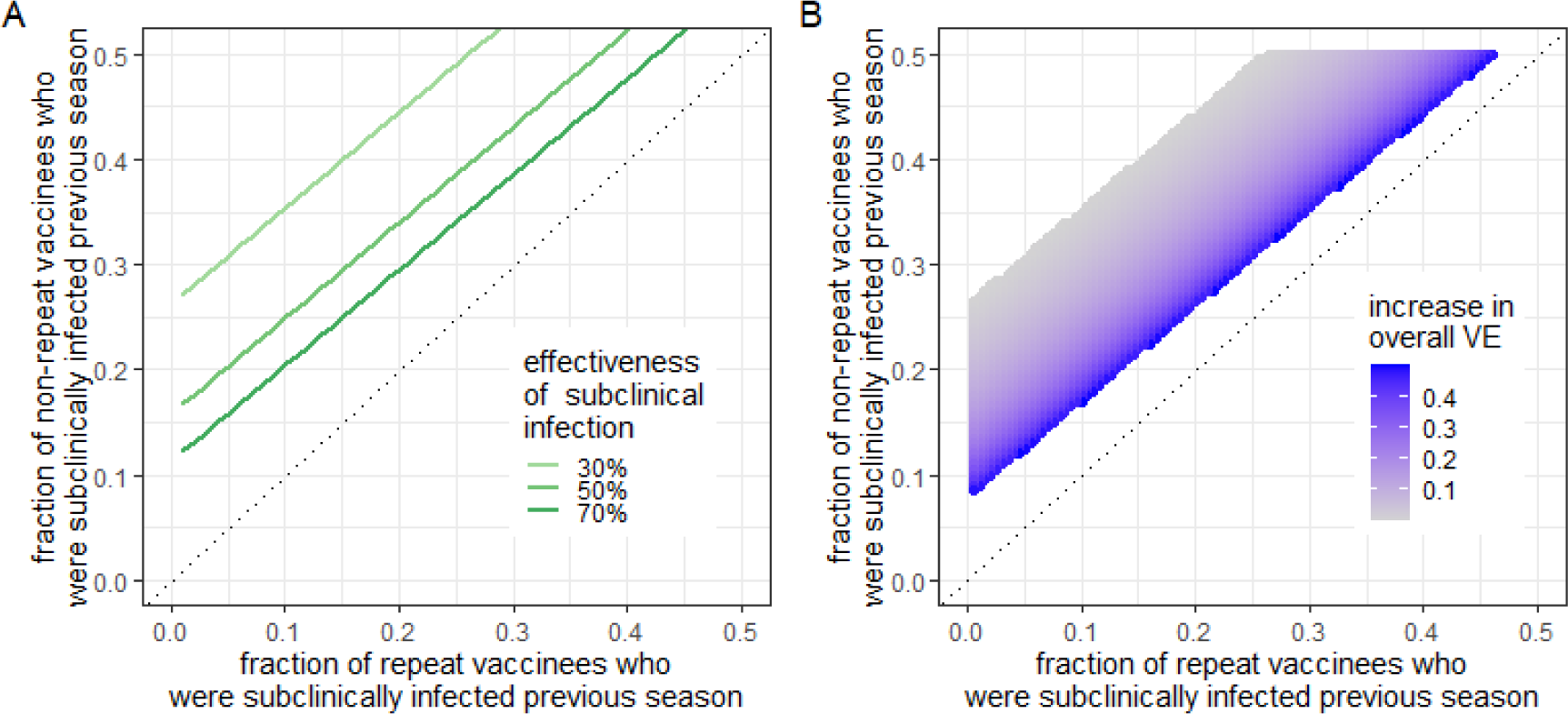
Subclinical infection might be able to explain the effect of repeated vaccination, aligning with the hypotheses of infection block (A) and enhanced immunogenicity (B) A) The fraction of repeat and non-repeat vaccinees who would need to have been subclinically infected in the previous season to reproduce the estimated effect of repeated vaccination in the US Flu VE Network (i.e., OR=1.1), given various assumptions of predetermined protection against clinical infection after subclinical infection (i.e., 30%, 50%, 70%). See Supplemental Section 5 for detailed methods. Under this hypothesis, subclinical infection is more common among those unvaccinated in the previous season (green lines above the 45-degree line) and reduces risk of infection this season. Only the plausible range of subclinical attack rate among repeat vaccinees (x-axis) and non-repeat vaccinees (y-axis; 0-50%) are shown in the figure. B) The absolute increase in VE (shown in the legend) from a baseline VE of 50% needed to reproduce the estimated effect of repeated vaccination in the US Flu VE Network (OR=1.1). Under this hypothesis, vaccination this season is more effective in those infected subclinically in the previous season, who are (as in A) more common among those unvaccinated in the previous season. The figure shows the scenario where the effectiveness of subclinical infection against future clinical infection is 30%. The uncolored portion of the figure represents the population where a boost in VE after infection will not generate the estimated effect of repeated vaccination (OR of 1.1). The results in both panels assume that vaccine effectiveness against clinical infection is 50%; clinical attack rate among vaccinees in a season is 1%; current-season clinical attack rate among the subset of current-season vaccinees not infected in the previous season is 1.5%; and clinical infection in the previous season perfectly protects against clinical infection in the following season.

For thoroughness, we showed that prior-season clinical infection is unlikely to be an important mediator in this relationship between prior-season vaccination and the odds of current-season clinical infection (Supplemental Section 6; SFig 6.1).

Compared with estimates from a low-clinical-incidence setting, in a high-incidence setting, we expect a greater excess of clinical infections in the current season among repeat vaccinees compared with non-repeat vaccinees (SFig 5.2).

#### 4.3.2 Enhanced vaccine immunogenicity hypothesis

If recent infection improves vaccine immunogenicity and thus vaccine-induced protection, a smaller difference in rates of subclinical infection between repeat and non-repeat vaccinees would generate the same estimated effect of repeated vaccination against infection in the current season (SFig 5.3). For example, in the scenario described in the second paragraph of section 4.3.1, we would observe the expected effect of repeated vaccination (OR=1.1) when the difference in the rate of subclinical infection between repeat and non-repeat vaccinees is 10% (e.g., ∼5% and ∼15% respectively), assuming subclinical infection reduces the probability of future clinical infection by 70%. But if recent infection boosts VE from 50% to 76%, a smaller difference in subclinical attack rates (∼5% in repeat vaccinees and ∼12% in non-repeat vaccinees) and weaker protection from subclinical infection (from 70% to 30%) can produce the same estimated effect.

## 5 Discussion

Observational studies[7–13,15,16], mostly using the test-negative design[8–13,15,16], have provided critical information on influenza VE. These studies can have biases and uncontrolled confounding that affect inference, including inference of VE in different subpopulations[33,39–42]. Reduced VE in repeat vaccinees has been a troubling, intermittent, and largely unexplained phenomenon[8,9]. We studied a component phenomenon, which is that the absolute risk (or odds) of infection among vaccinees in the current season is less if they were unvaccinated last season than if they were vaccinated last season. We observed waning of vaccine protection and repeat vaccinees’ tendency to vaccinate earlier within a season compared with non-repeat vaccinees. We showed that clinical infection impacts individuals’ decisions to vaccinate in the next season, and clinical infections in the past 1-2 seasons strongly protect against reinfection. However, these potentially biasing factors – prior-season infection and timing of vaccination – could not fully explain the higher risk of infection in the repeat vaccinees vs. non-repeat vaccinees in our study population. We showed that the residual repeat vaccination effect might be explained by different rates of subclinical infection between repeat and non-repeat vaccinees via two proposed mechanisms, the infection block hypothesis[4,24-26] and enhanced vaccine immunogenicity and protection post-infection[28,43]. The difference in rates of subclinical infection between the two groups and its variation from one season to the next might thus underlie variability in estimated effects of repeated vaccination.

Mostly due to lack of data, clinical infection history has typically not been accounted for when estimating influenza VE. We found that accounting for clinical infection history did not substantially change the estimated effect of repeat vaccination, indicating that confounding by prior-season clinical infection may not fully explain the elevated odds of infection among repeat vaccinees. Aside from its potential role as a confounder, we found clinical infection unlikely to act as an important mediator. Verifying the finding in surveillance data requires methods that can tease apart the direct and indirect effect of vaccination after taking into account the interaction of vaccination and infection over a multi-year period.

The sensitivity of the estimated effect of repeated vaccination to differences in subclinical attack rates and infection-associated protection suggests a possible explanation for the observed variability in the estimated effect of repeat vaccination and other VE measures across locations and time. There is well-known spatiotemporal variation in the sizes of influenza epidemics and in circulating clades that could affect the amount of protection conferred by infection in different populations. Our results suggest a need to try to account more precisely for past infections, so that VE estimates can be compared across populations stratified by similar infection history. Longitudinal cohort studies that involve blood collection, active surveillance, and sequencing can be useful for identifying subclinical infections, and coupling these observations with healthcare-seeking behavior and PCR testing can help test the infection block and enhanced immunogenicity hypotheses[44]. Eventually, stratification on infection history may be possible through surrogate immune markers.

The study has several limitations. Throughout our analysis, we assumed that influenza vaccination with any type of influenza vaccine confers complete protection in a subset of vaccinees. We did not consider “leaky” vaccine effects, where vaccines are partially protective in all recipients, and which can lead to an observed decline in VE estimates even when vaccine protection does not wane [45]. Although the test-negative study, by selecting only patients who seek medical care, is designed to reduce the difference in health-seeking behavior between cases and non-cases, it does not eliminate it[40]. We did not explore birth cohort effects or the effects of antigenic distance on protection[46–49]. Since we assumed individuals without an enrollment record in the previous season were not clinically infected, some of them may have been misclassified.

The practical benefits of annual vaccination programs should not be extrapolated from this analysis of the relative risk of infection in repeat vaccinees compared to non-repeat vaccinees. The choice between an annual and a non-annual vaccination program should be based on assessments of the infection risk among all repeat and non-repeat vaccinees as well as the unvaccinated. Our analysis does not compare the risk of infection between repeat vaccinees and those vaccinated in the prior season only, who would be part of a hypothetical non-annual vaccination program.

Our study provides evidence that two potential factors, timing of vaccination and clinical infection history, cannot fully explain the increased infection risk in repeat vaccinees compared with non-repeat vaccinees. Clinical infection history is further unlikely to act as a strong mediator to explain the repeated vaccination effect. Instead, under reasonable assumptions, the infection block and enhanced-immunogenicity hypotheses involving subclinical infection in the previous season may explain the effect, thus acting as a potential mediator. Estimation of VE requires careful consideration of both time since vaccination and infection history of different subpopulations.

## Data Availability

Data and relevant code are available at https://github.com/cobeylab/FluVE_repeatvac_public

## Acknowledgements

QB, BAD, ML, SC conceived of the study. HQM, ETM, MG, KJW, BG, BF collected the data. QB performed the analyses and generated all figures. QB, BAD, ML, SC wrote the manuscript. All authors revised the manuscript. We thank Benjamin A. Cowling for comments. Authors report no conflict of interest.

Investigators and collaborators in the US Flu VE Network Investigators include:

Baylor Scott & White Health – Kempapura Murthy, Chandni Raiyani, Kayan Dunnigan, Muffadal Mamawala

CDC – Jessie R. Chung, Manish Patel

Henry Ford Medical Center – Lois Lamerato

Kaiser Permanente Washington – Michael L. Jackson, C. Hallie Phillips, Erika Kiniry

Marshfield Clinic Research Institute – Edward A. Belongia, Jennifer P. King

University of Michigan – Arnold S. Monto

University of Pittsburgh – Richard K. Zimmerman, Mary Patricia Nowalk, Krissy Moehling Geffel

## Footnotes

1 The authors report no conflict of interest; This study was partially supported by the Collaborative Influenza Vaccine Innovation Centers (CIVICs) contract 75N93019C00051 (S.C. and Q.B.). ML acknowledges support from US NIH grant 1R01GM139926 to University of Michigan. Corresponding author’s contact information. Alternative corresponding author: Sarah Cobey. Contact information of the alternative corresponding author. Data and relevant code are available at https://github.com/cobeylab/FluVE_repeatvac_public. The analyses were conducted in R version 4.1.1.

## References

1. Vaccines against influenza WHO position paper – November 2012. Wkly Epidemiol Rec. 2012; 87(47):461–476.

3. Principi N, Camilloni B, Esposito S, ESCMID Vaccine Study Group (EVASG). Influenza immunization policies: Which could be the main reasons for differences among countries? Hum Vaccin Immunother. 2018; 14(3):684–692.

3. Hoskins TW, Davies J, Smith AJ, Allchin A, Miller C, Pollock TM. INFLUENZA AT CHRIST’S HOSPITAL: MARCH, 1974 [Internet]. The Lancet. 1976. p. 105–108. Available from: 10.1016/s0140-6736(76)93151-2

4. Hoskins TW, Davies JR, Smith AJ, Miller CL, Allchin A. Assessment of inactivated influenza-A vaccine after three outbreaks of influenza A at Christ’s Hospital. Lancet. 1979; 1(8106):33–35.

5. Skowronski DM, Janjua NZ, De Serres G, et al. A Sentinel Platform to Evaluate Influenza Vaccine Effectiveness and New Variant Circulation, Canada 2010–2011 Season [Internet]. Clinical Infectious Diseases. 2012. p. 332–342. Available from: 10.1093/cid/cis431

6. Ng S, Fang VJ, Ip DKM, et al. Humoral antibody response after receipt of inactivated seasonal influenza vaccinations one year apart in children. Pediatr Infect Dis J. 2012; 31(9):964–969.

7. Ohmit SE, Petrie JG, Malosh RE, et al. Influenza Vaccine Effectiveness in the Community and the Household [Internet]. Clinical Infectious Diseases. 2013. p. 1363–1369. Available from: 10.1093/cid/cit060

8. Belongia EA, Skowronski DM, McLean HQ, Chambers C, Sundaram ME, De Serres G. Repeated annual influenza vaccination and vaccine effectiveness: review of evidence. Expert Rev Vaccines. 2017; 16(7):1–14.

9. Ramsay LC, Buchan SA, Stirling RG, et al. The impact of repeated vaccination on influenza vaccine effectiveness: a systematic review and meta-analysis. BMC Med. 2019; 17(1):9.

10. McLean HQ, Thompson MG, Sundaram ME, et al. Impact of Repeated Vaccination on Vaccine Effectiveness Against Influenza A(H3N2) and B During 8 Seasons [Internet]. Clinical Infectious Diseases. 2014. p. 1375–1385. Available from: 10.1093/cid/ciu680

11. Ohmit SE, Thompson MG, Petrie JG, et al. Influenza Vaccine Effectiveness in the 2011–2012 Season: Protection Against Each Circulating Virus and the Effect of Prior Vaccination on Estimates [Internet]. Clinical Infectious Diseases. 2014. p. 319–327. Available from: 10.1093/cid/cit736

12. Skowronski DM, Chambers C, Sabaiduc S, et al. A Perfect Storm: Impact of Genomic Variation and Serial Vaccination on Low Influenza Vaccine Effectiveness During the 2014-2015 Season. Clin Infect Dis. 2016; 63(1):21–32.

13. Valenciano M, Kissling E, Reuss A, et al. Vaccine effectiveness in preventing laboratory-confirmed influenza in primary care patients in a season of co-circulation of influenza A(H1N1)pdm09, B and drifted A(H3N2), I-MOVE Multicentre Case-Control Study, Europe 2014/15. Euro Surveill. 2016; 21(7):ii=30139.

14. Kissling E, Pozo F, Buda S, et al. Low 2018/19 vaccine effectiveness against influenza A(H3N2) among 15-64-year-olds in Europe: exploration by birth cohort. Euro Surveill [Internet]. 2019; 24(48). Available from: 10.2807/1560-7917.ES.2019.24.48.1900604

15. McLean HQ, Thompson MG, Sundaram ME, et al. Influenza vaccine effectiveness in the United States during 2012-2013: variable protection by age and virus type. J Infect Dis. 2015; 211(10):1529–1540.

16. Kissling E, Pozo F, Buda S, et al. Low 2018/19 vaccine effectiveness against influenza A(H3N2) among 15–64-year-olds in Europe: exploration by birth cohort [Internet]. Eurosurveillance. 2019. Available from: 10.2807/1560-7917.es.2019.24.48.1900604

17. Ferdinands JM, Shay DK. Magnitude of potential biases in a simulated case-control study of the effectiveness of influenza vaccination. Clin Infect Dis. 2012; 54(1):25–32.

18. Ray GT, Lewis N, Klein NP, et al. Intraseason Waning of Influenza Vaccine Effectiveness. Clin Infect Dis. 2019; 68(10):1623–1630.

19. Ferdinands JM, Gaglani M, Martin ET, et al. Waning Vaccine Effectiveness Against Influenza-Associated Hospitalizations Among Adults, 2015–2016 to 2018–2019, United States Hospitalized Adult Influenza Vaccine Effectiveness Network [Internet]. Clinical Infectious Diseases. 2021. p. 726–729. Available from: 10.1093/cid/ciab045

30. Young B, Sadarangani S, Jiang L, Wilder-Smith A, Chen MI-C. Duration of Influenza Vaccine Effectiveness: A Systematic Review, Meta-analysis, and Meta-regression of Test-Negative Design Case-Control Studies. J Infect Dis. 2018; 217(5):731–741.

31. Jiménez-Jorge S, Mateo S de, Delgado-Sanz C, et al. Effectiveness of influenza vaccine against laboratory-confirmed influenza, in the late 2011-2012 season in Spain, among population targeted for vaccination. BMC Infect Dis. 2013; 13:441.

32. Ferdinands JM, Fry AM, Reynolds S, et al. Intraseason waning of influenza vaccine protection: Evidence from the US Influenza Vaccine Effectiveness Network, 2011-12 through 2014-15. Clin Infect Dis. 2017; 64(5):544–550.

33. Fox SJ, Miller JC, Meyers LA (2017) Seasonality in risk of pandemic influenza emergence. PLoS Comput Biol 13(10): e1005749.

34. Skowronski DM, Janjua NZ, Hottes TS, De Serres G. Mechanism for Seasonal Vaccine Effect on Pandemic H1N1 Risk Remains Uncertain. Clin. Infect. Dis. 2011. p. 831–2; author reply 832–3.

35. Skowronski DM, De Serres G, Crowcroft NS, et al. Association between the 2008-09 seasonal influenza vaccine and pandemic H1N1 illness during Spring-Summer 2009: four observational studies from Canada. PLoS Med. 2010; 7(4):e1000258.

36. Cowling BJ, Ng S, Ma ESK, et al. Protective Efficacy of Seasonal Influenza Vaccination against Seasonal and Pandemic Influenza Virus Infection during 2009 in Hong Kong [Internet]. Clinical Infectious Diseases. 2010. p. 1370–1379. Available from: 10.1086/657311

37. Ranjeva, S., Subramanian, R., Fang, V.J. et al. Age-specific differences in the dynamics of protective immunity to influenza. Nat Commun 10, 1660 (2019). 10.1038/s41467-019-09652-6

38. Auladell M, Phuong HVM, Mai LTQ, et al. Influenza virus infection history shapes antibody responses to influenza vaccination. Nat Med. 2022; 28(2):363–372.

39. Krammer F. The human antibody response to influenza A virus infection and vaccination. Nat Rev Immunol. 2019; 19(6):383–397.

30. Hall V, Foulkes S, Insalata F, et al. Protection against SARS-CoV-2 after Covid-19 Vaccination and Previous Infection. N Engl J Med. 2022; 386(13):1207–1220.

31. VanderWeele TJ. Explanation in causal inference: developments in mediation and interaction [Internet]. International Journal of Epidemiology. 2016. p. dyw277. Available from: 10.1093/ije/dyw277

32. Hernan MA, Robins JM. Causal Inference. CRC Press; 2019.

33. Gaglani M, Pruszynski J, Murthy K, et al. Influenza Vaccine Effectiveness Against 2009 Pandemic Influenza A(H1N1) Virus Differed by Vaccine Type During 2013–2014 in the United States [Internet]. Journal of Infectious Diseases. 2016. p. 1546–1556. Available from: 10.1093/infdis/jiv577

34. Zimmerman RK, Nowalk MP, Chung J, et al. 2014-2015 Influenza Vaccine Effectiveness in the United States by Vaccine Type. Clin Infect Dis. 2016 Dec 15;63(12):1564–1573. 10.1093/cid/ciw635.

35. Jackson ML, Chung JR, Jackson LA, et al. Influenza Vaccine Effectiveness in the United States during the 2015-2016 Season. N Engl J Med. 2017 Aug 10;377(6):534–543. 10.1056/NEJMoa1700153.

36. Kieke AL, Kieke BA Jr, Kopitzke SL, et al. Validation of Health Event Capture in the Marshfield Epidemiologic Study Area. Clin Med Res. 2015; 13(3-4):103–111.

37. Cohen C, Kleynhans J, Moyes J, et al. Asymptomatic transmission and high community burden of seasonal influenza in an urban and a rural community in South Africa, 2017–18 (PHIRST): a population cohort study. The Lancet Global Health. 2021; 9:e863–e874.

38. Jackson ML, Jackson LA, Kieke B, et al. Incidence of medically attended influenza infection and cases averted by vaccination, 2011/2012 and 2012/2013 influenza seasons [Internet]. Vaccine. 2015. p. 5181–5187. Available from: 10.1016/j.vaccine.2015.07.098

39. Lipsitch M, Tchetgen Tchetgen E, Cohen T. Negative controls: a tool for detecting confounding and bias in observational studies. Epidemiology. 2010; 21(3):383–388.

40. Sullivan SG, Tchetgen Tchetgen EJ, Cowling BJ. Theoretical Basis of the Test-Negative Study Design for Assessment of Influenza Vaccine Effectiveness. Am J Epidemiol. 2016; 184(5):345– 353.

41. Chua H, Feng S, Lewnard JA, et al. The Use of Test-negative Controls to Monitor Vaccine Effectiveness: A Systematic Review of Methodology. Epidemiology. 2020; 31(1):43–64.

42. Jackson ML, Nelson JC. The test-negative design for estimating influenza vaccine effectiveness [Internet]. Vaccine. 2013. p. 2165–2168. Available from: 10.1016/j.vaccine.2013.02.053

43. Altarawneh HN, Chemaitelly H, Ayoub HH, et al. Effects of Previous Infection and Vaccination on Symptomatic Omicron Infections. N Engl J Med. 2022; 387(1):21–34.

44. Monto AS, DeJonge PM, Callear AP, et al. Coronavirus Occurrence and Transmission Over 8 Years in the HIVE Cohort of Households in Michigan. J Infect Dis. 2020; 222(1):9–16.

45. Tokars JI, Patel MM, Foppa IM, Reed C, Fry AM, Ferdinands JM. Waning of Measured Influenza Vaccine Effectiveness Over Time: The Potential Contribution of Leaky Vaccine Effect. Clin Infect Dis. 2020; 71(10):e633–e641.

46. Smith DJ, Forrest S, Ackley DH, Perelson AS. Variable efficacy of repeated annual influenza vaccination. Proc Natl Acad Sci U S A. 1999; 96(24):14001–14006.

47. Arevalo P, McLean HQ, Belongia EA, Cobey S. Earliest infections predict the age distribution of seasonal influenza A cases. Elife [Internet]. 2020; 9. Available from: 10.7554/eLife.50060

48. Skowronski DM, Chambers C, De Serres G, et al. Serial Vaccination and the Antigenic Distance Hypothesis: Effects on Influenza Vaccine Effectiveness During A(H3N2) Epidemics in Canada, 2010-2011 to 2014-2015. J Infect Dis. 2017; 215(7):1059–1099.

49. Flannery B, Smith C, Garten RJ, et al. Influence of Birth Cohort on Effectiveness of 2015-2016 Influenza Vaccine Against Medically Attended Illness Due to 2009 Pandemic Influenza A(H1N1) Virus in the United States. J Infect Dis. 2018; 218(2):189–196.

