## Supplemental material for "Reduced effectiveness of repeat influenza vaccination: distinguishing among within-season waning, recent clinical infection, and subclinical infection"

**Study group team members are listed in the acknowledgements

*Equal contribution

6 Supplemental Material

Section 1. Summary of study participants, study enrollment, and influenza seasonality

Additional information on study setting and study population: In this study, repeat vaccinees were defined as individuals vaccinated in both the current season and the previous season. Non-repeat vaccinees were defined as individuals vaccinated in the current season only. Conditions that may increase the risk of complications attributable to severe influenza are defined as high-risk conditions. High-risk conditions were chronic pulmonary (including asthma), cardiovascular (excluding isolated hypertension), renal, hepatic, neurologic, hematologic, or metabolic disorders (including diabetes mellitus). Persons who were immunocompromised due to any cause, including but not limited to immunosuppression caused by medications or human immunodeficiency virus infection, were also considered to have a high-risk condition.

Surveillance data from the MCHS were available between the 2007-2008 season and the 2018-2019 season. In total, 16,378 individuals were enrolled during these 12 seasons and contributed 24,399 visits (Supplementary Figure 1.1). Among them, 15,459 individuals were included in the main analyses that study the impact of clinical infection (Supplementary Table 1.2). These individuals contributed 21,188 visits. Among the 7,390 individuals who had received one dose of the current seasonal influenza vaccine ≥14 days prior to illness onset, 78.4% (n=7,362) had also been vaccinated in the previous season, and 90.9% (n=8,539) had been vaccinated in at least one of the three seasons immediately before the enrollment season. Among those who presented with acute respiratory symptoms and were eligible for enrollment in the prior season, 71.2% (1,002/1,407) were enrolled.

Table 1.1: Characteristics of the vaccinated individuals in the US Flu VE Network by their influenza infection status in the current season and vaccination status in the prior season, from the 2011-2012 through the 2018-2019 seasons.

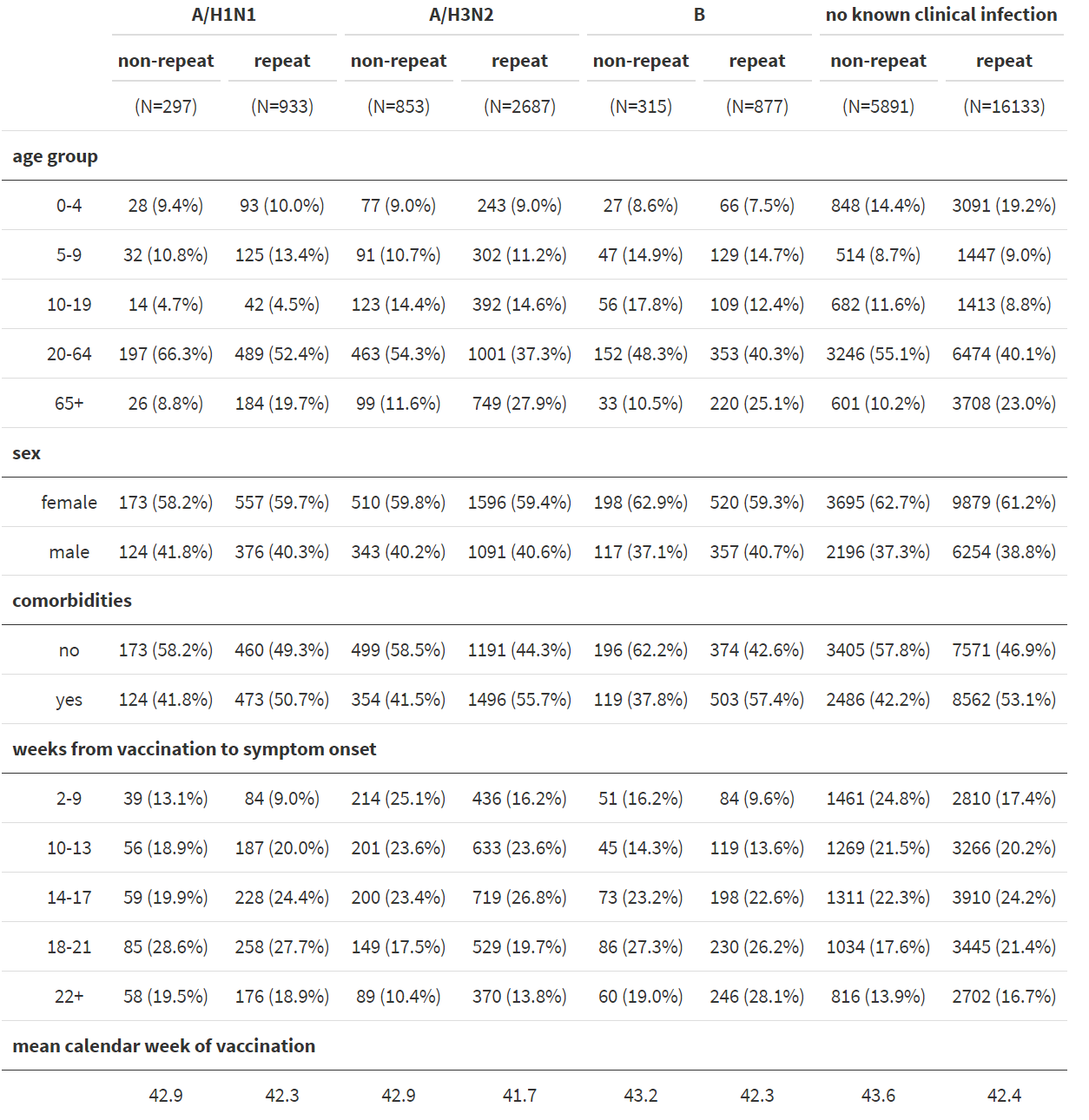

Table 1.2: Characteristics of the vaccinated individuals at the Marshfield Clinic Health System by their influenza infection status in the current season and vaccination status in the previous season, from the 2008-2009 through the 2017-2018 seasons.

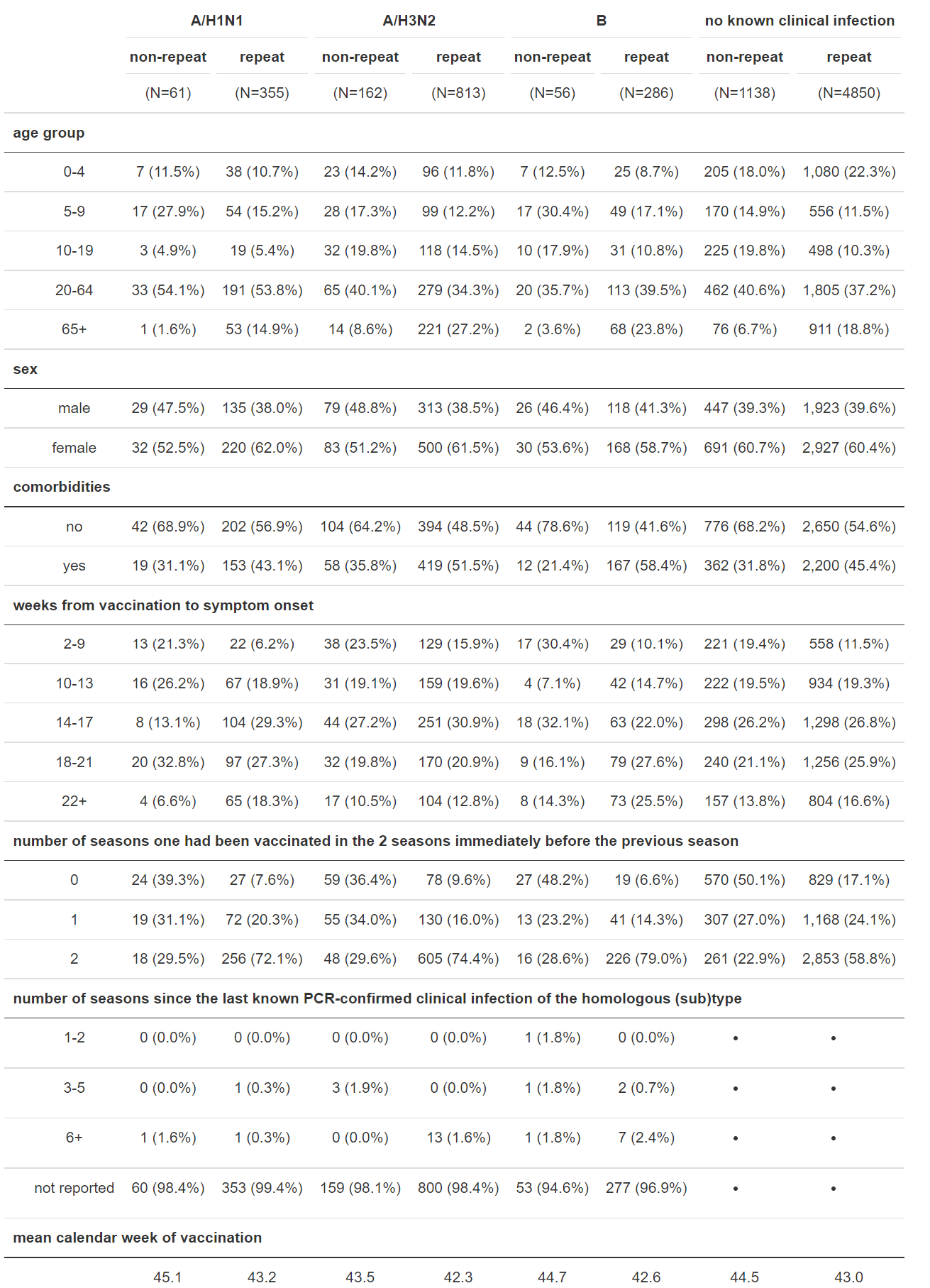

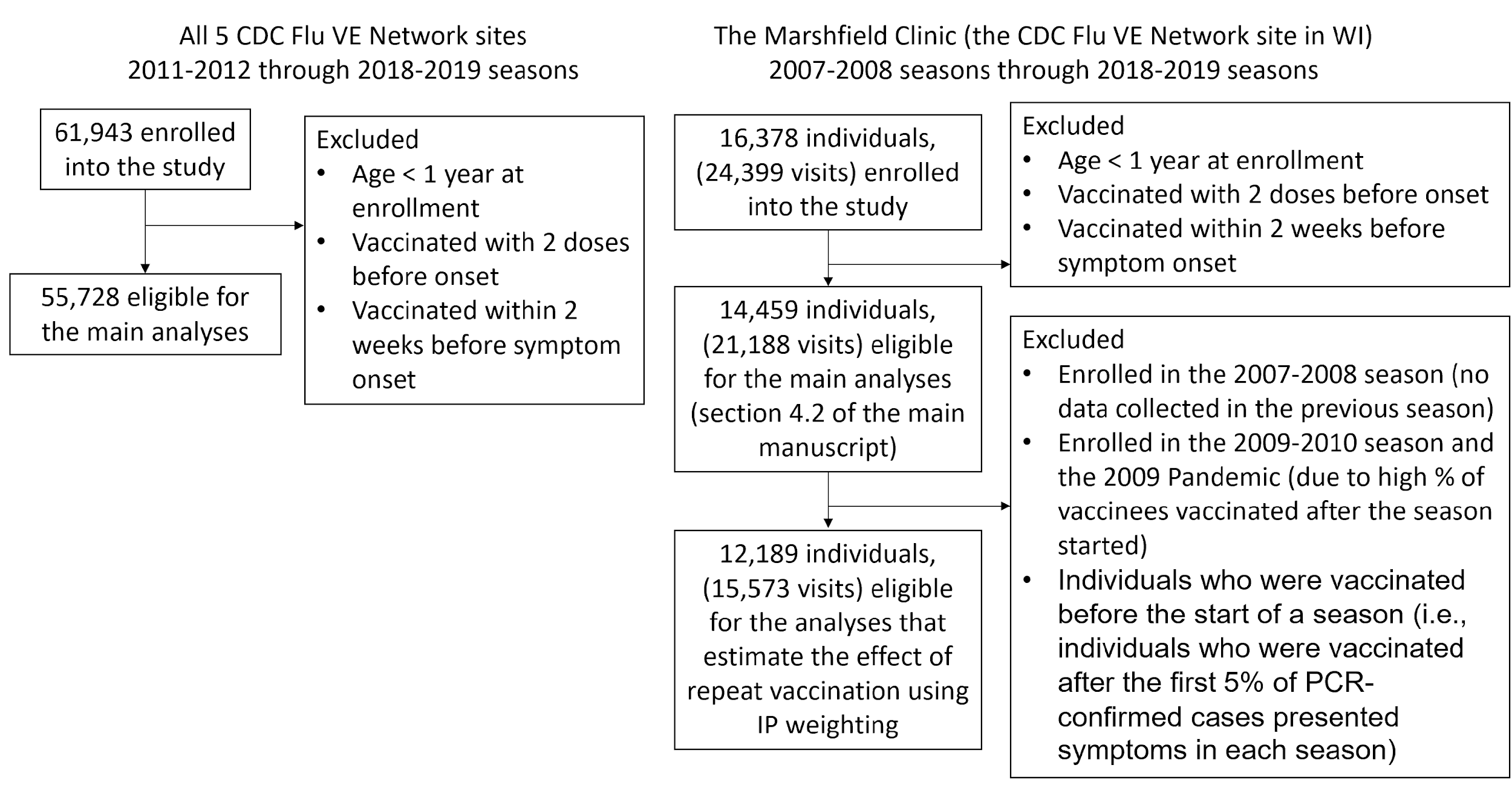

Figure 1.1: Enrollment of study participants and exclusion criteria

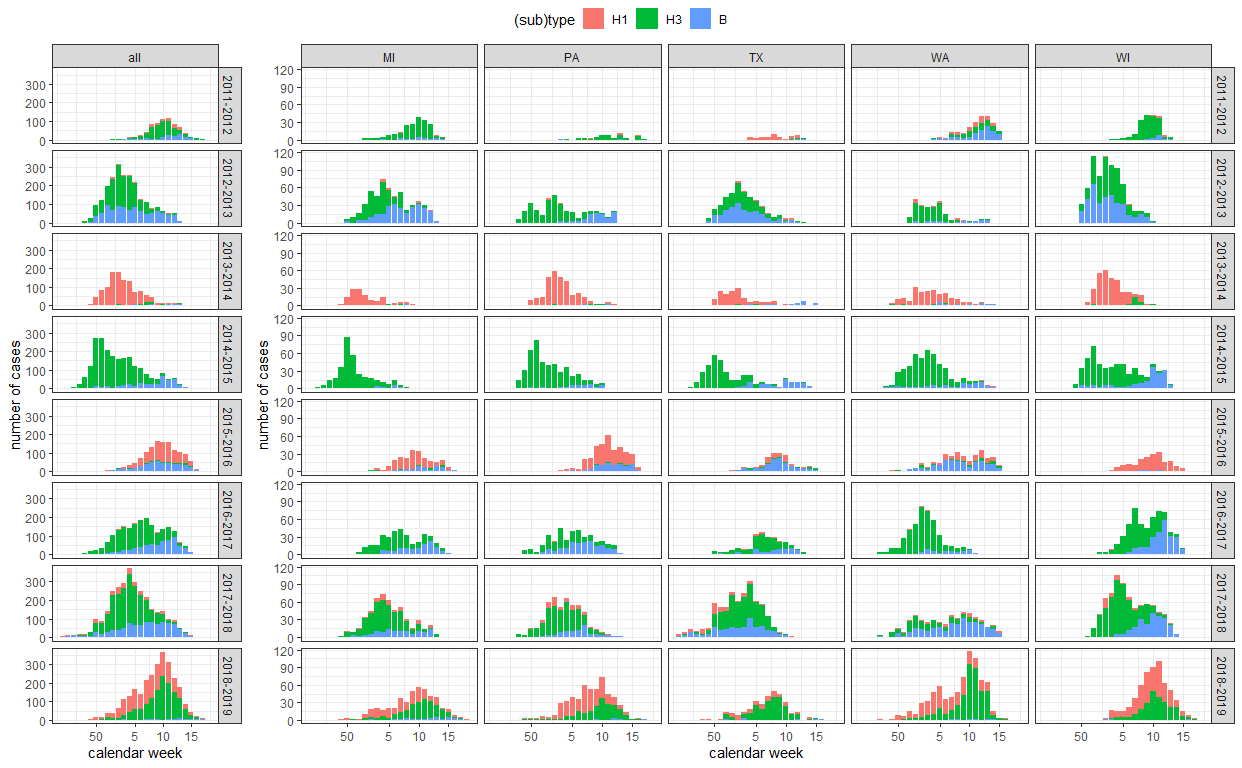

Figure 1.2: Seasonality by influenza (sub)type over the study period across all 5 US Flu VE Network sites

Section 2. Conceptual framework for studying the effect of repeat vaccination

The effect of repeated vaccination can be conceptualized as a contrast of vaccination strategies sustained over two influenza seasons (the previous and the current season). That is, the repeat vaccine strategy involves receipt of an influenza vaccine in the previous season and current season, and the non-repeat vaccine strategy involves no receipt of an influenza vaccine in the previous season but receipt of one in the current season. We include a directed acyclic graph (Supplementary Figure 2.1) that encodes our assumptions about the underlying causal structure in this two-season framework, and we describe the potential confounding and selection bias [[51]](https://paperpile.com/c/kjBGeu/P1GL). However, due to data constraints, we simplified parts of the analyses to address questions under a single-season framework.

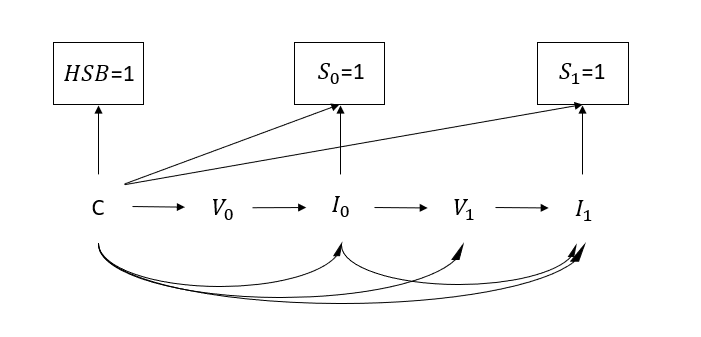

Figure 2.1: A directed acyclic graph that encodes our assumptions about the underlying causal relationships of vaccination and infection status in a test-negative design that spans two seasons. *V* denotes vaccination status. *I* denotes infection status. *S* denotes selection into the study. *C* denotes baseline confounders. *HSB* denotes healthcare-seeking behavior. Subscripts 0 and 1 represent status at the previous and current seasons, respectively.

Underlying causal structure: Estimating the effect of repeated vaccination can be conceptualized as the estimation of the joint effect of vaccination in the previous season (*V_0_*) and the current season (*V_1_*) on infection outcome in the current season (*I_1_*). We denote vaccination status by *V* and infection status by *I*. Subscripts 0 and 1 represent status at the previous and current seasons, respectively. The arrows V*_0_* → *I_0_* and *V_1_* → *I_1_* represent the effect of vaccination on infection in each season. *I_0_* →*I_1_* represents the effect of infection in the previous season on infection in the current season, and *I_0_* →*V_1_* represents the effect of influenza infection in the previous season on the decision to vaccinate in the next season, the evidence for which is presented in section 4.2 in the main text.

Potential confounding and selection bias: Selection into the test-negative design in each season, denoted by *S_0_* and *S_1_*, is a result of an individual experiencing acute respiratory illness, seeking care at ambulatory facilities, and getting tested for influenza infection, and this chain of events is denoted by $I_{k}$ → $S_{k}$ (*k ϵ* {0,1}). Healthcare-seeking behavior, denoted by *HSB*, may affect $S_{k}$, $V_{k}$, and $I_{k}$ because subjects with healthcare-seeking proclivities may be more likely to seek care, vaccinate, and practice healthier behaviors that reduce the odds of infection. Other clinical or demographic factors, such as age, sex, and comorbidities, denoted by confounders *C*, can also affect $S_{k}$, $V_{k}$, and $I_{k}$. The test-negative design assumes that by restricting recruitment to those who seek health care, the study subjects have identical healthcare seeking behavior (*HSB*=1), thus reducing confounding [[51]](https://paperpile.com/c/kjBGeu/P1GL). In the present application, we assume no unmeasured confounding given this restriction and further adjustment for measured variables.

Accounting for within-season waning of vaccine protection: For the analyses in this section, because infection status in the previous season was not available across all sites in the US Flu VE Network, we estimated relative odds of infection under a single-season framework and treated previous-season vaccination status as a pre-baseline variable (a component of the *C* node in Supplementary Figure 2.1).

Adjustment for clinical infection history: We first determined how a clinical infection outcome in the current season is associated with clinical infection with the homologous (sub)types and separately with the heterologous (sub)types in any prior season (the *I_0_*→*I_1_* edge in Supplementary Figure 2.1). Next, we assessed whether clinical influenza virus infections in the previous season influenced the decision to vaccinate in the current season using logistic regression models (the *I_0_*→*V_1_* edge in Supplementary Figure 2.1). We then estimated the effect of repeated vaccination after adjusting for clinical infection history under a two-season framework.

Impact of subclinical infection history: In this section, fundamentally, we aimed to assess the impact of misclassification of infection status in the previous season (the *I_0_* node in Supplementary Figure 2.1) using a single-season framework.

Section 3. Models that adjust and do not adjust for waning vaccination protection

3.1 Detailed methods

For the analyses in this section, we used data across the five sites in the US Flu VE Network. We first determined whether the timing of vaccination in the current season differed between repeat and non-repeat vaccinees. We restricted this analysis to current-season vaccinees and fit a linear regression model with the dependent variable of calendar week of vaccination in the current season and the following independent variables: prior-season vaccination status, age group (0-4, 5-9, 10-19, 20-64, 65+), sex, presence of at least one documented high-risk condition in the prior season (referred to as comorbidity), and influenza season.

Using logistic regression models, we then estimated the relative odds of clinical infection among repeat vaccinees with reference to non-repeat vaccinees after adjusting for time of vaccination in the current season to account for the waning of vaccine protection. Clinical infection status in the current season when waning vaccine effectiveness is adjusted for is modeled as

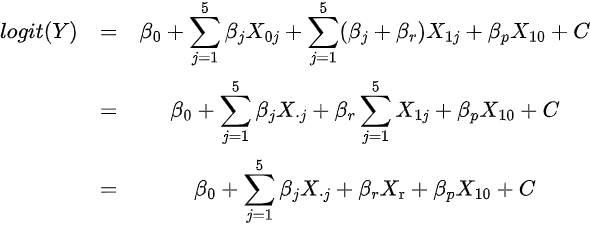

, where Y is the probability of clinical infection in the current season; *X_ij_* is an indicator for vaccination status in the prior season (*i*=0 if not vaccinated in the prior season, 1 if vaccinated, · if either vaccinated or not vaccinated) and the timing of vaccination in the current season (*j*=0 if not vaccinated in the current season, 1 if vaccinated 2-9 weeks after vaccination, 2 if 10-13 weeks after, 3 if 14-17 weeks after, 4 if 18-21 weeks after, and 5 if 22+ weeks after). *X_r_* represents repeat vaccinees. *C* represents other baseline variables adjusted in the model (i.e., age group, sex, comorbidities, influenza season, study site, and calendar month of symptom onset). Then, $e^{\beta_{j}}$ is the OR for clinical infection in the *j^th^* interval after current-season vaccination compared with those not vaccinated in either season. $e^{\beta_{r}}$ is the OR for clinical infection among repeat vaccinees compared with non-repeat vaccinees who were only vaccinated in the current season. $e^{\beta_{p}}$ is the OR for clinical infection among those vaccinated in the prior season but not this season compared with those not vaccinated in either season (Figure 1B).

Clinical infection status in the current season when waning vaccine effectiveness is not adjusted for is modeled as follows:

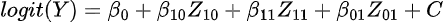

where *Z_ik_* is an indicator for vaccination status in the previous season (i=1 vs. 0) and in the current season (k=1 vs. 0). $e^{\beta11-\beta01}$ is the OR for clinical infection among repeat vaccinees compared with current-season vaccinees who were not vaccinated in the previous season. The results generated by these models are shown in section 4.1 and Figure 1B-C in the main text.

3.2 Additional results

After adjusting for the timing of vaccination in the current season, we found that repeat vaccinees had 1.03 (95%CI: 0.89-1.18) times the odds of testing positive for type B than non-repeat vaccinees, similar to the OR of 1.06 (95%CI: 0.92-1.22) before adjustment (Figure 1C). Likewise, the adjustment did not strongly alter the odds of clinical infection with A/H1N1pdm09 among repeat vaccinees compared with non-repeat vaccinees (pre-adjustment OR=1.08 (95%CI: 0.94-1.24) to post-adjustment OR=1.03 (95%CI: 0.90-1.19)). The adjustment did not change the observation that repeat vaccinees had higher odds of clinical infection with A/H3N2 than non-repeat vaccinees (post-adjustment OR=1.11 (95%CI: 1.02-1.21) vs. pre-adjustment OR=1.13 (95%CI: 1.04-1.23)). The effect of repeated vaccination against A/H3N2 was particularly strong among the 10-19-year-olds (post-adjustment OR=1.48, 95%CI: 1.18, 1.87; Supplementary Figure 2.2). In summary, adjusting for the timing of vaccination in the current season did not notably change the findings of a marked repeat vaccination effect for A/H3N2 and had little to no effect for A/H1N1pdm09 and type B.

Compared with individuals not vaccinated in either season, the odds of testing positive for infection with type B increased from 0.37 (95%CI: 0.30-0.45) among individuals vaccinated 2-9 weeks before testing to 0.54 (95%CI: 0.46, 0.64) 18-21 weeks before testing. Compared with individuals not vaccinated in either season, the odds of testing positive for infection with A/H3N2 increased from 0.68 (95%CI: 0.61-0.76) among individuals vaccinated 2-9 weeks before testing to 0.84 (95%CI: 0.75-0.94) 18-21 weeks before testing.

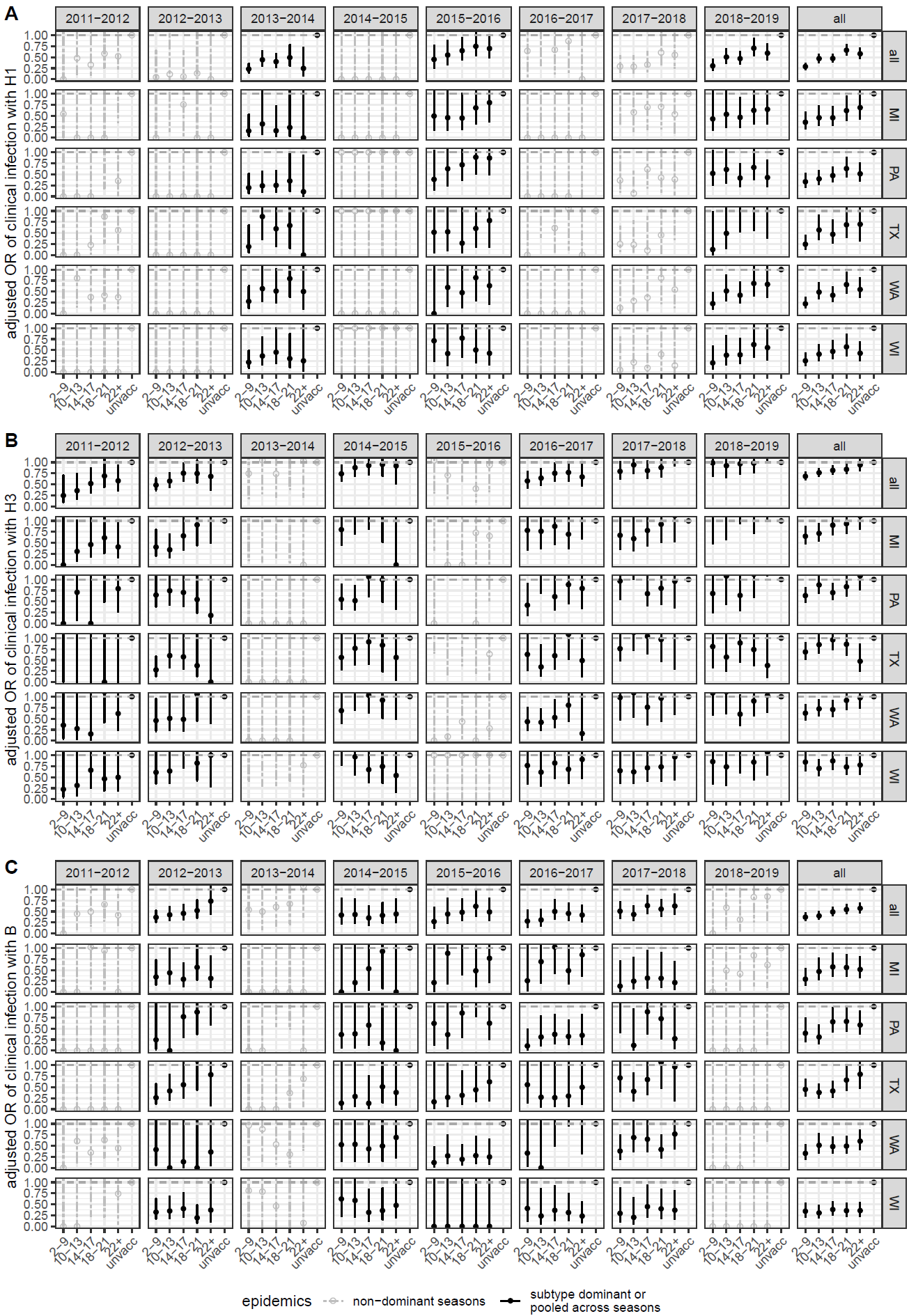

Figure 3.1: Waning vaccine protection by subtype, season, and site. Panels A-C represent the adjusted odds ratio for clinical infection with A/H1N1pdm09 (A), A/H3N2 (B), and type B (C) comparing individuals tested 2-9, 10-13, 14-17, 18-21, and 22+ weeks after vaccination with respect to those not vaccinated in the current season. The solid lines represent relative odds in seasons when there was significant circulation of the (sub)type. (Sub)type composition of PCR-confirmed influenza infection across all 5 Flu VE Network sites is shown in Supplementary Figure 1.2. In all panels, error bars indicate 95% confidence intervals.

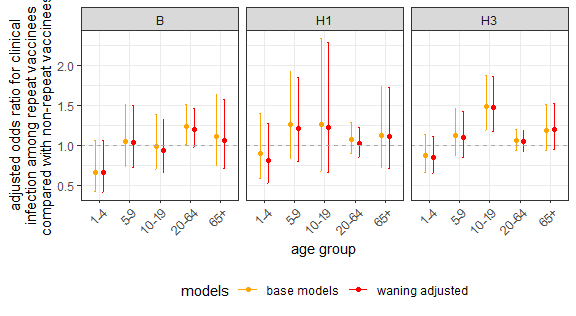

Figure 3.2: Estimated effect of repeated vaccination by age group across all 5 study sites from the 2011-2012 through the 2018-2019 seasons. Error bars indicate 95% confidence intervals.

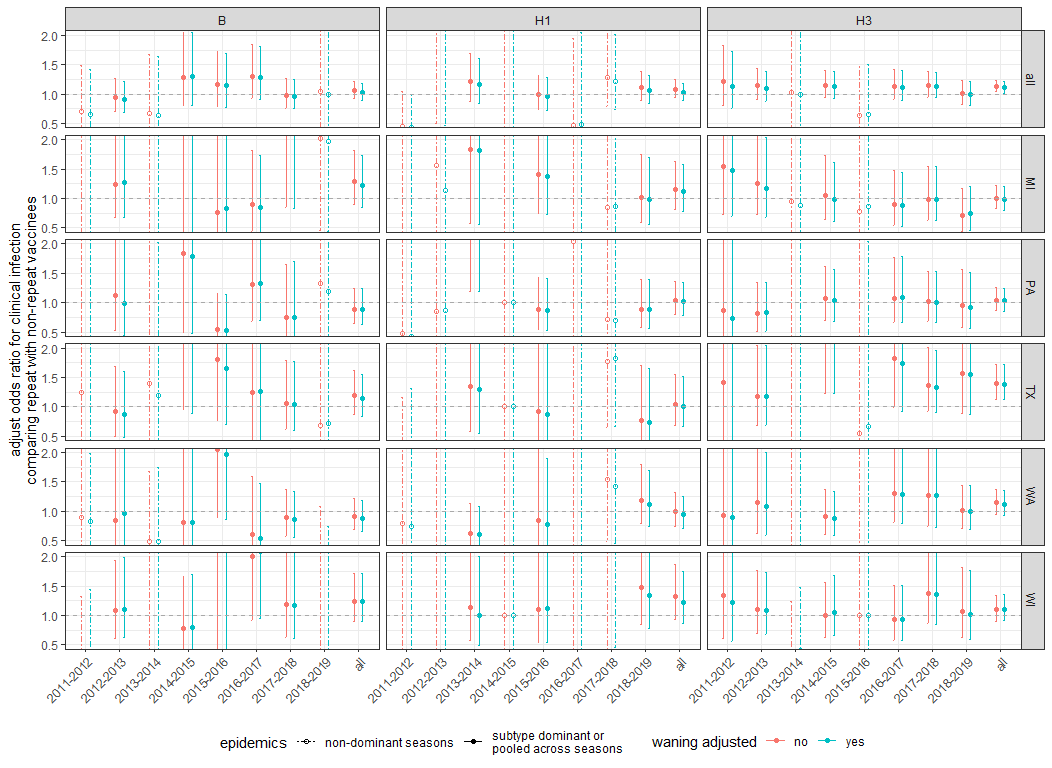

Figure 3.3: Impact of the timing of vaccination in the current season and waning of vaccine-induced immunity. The figure shows the adjusted odds ratio for clinical infection comparing repeat vaccinees with non-repeat vaccinees before (red) and after (blue) adjusting for timing of vaccination by season and site. The solid lines represent estimates in seasons when A/H3N2 was the dominant subtype, when A/H1N1pdm09 and A/H3N2 cocirculated, or there was significant circulation of type B. (Sub)type composition of PCR-confirmed influenza infections across all 5 sites in the US Flu VE Network are shown in Supplementary Figure 1.2. In all panels, error bars indicate 95% confidence intervals.

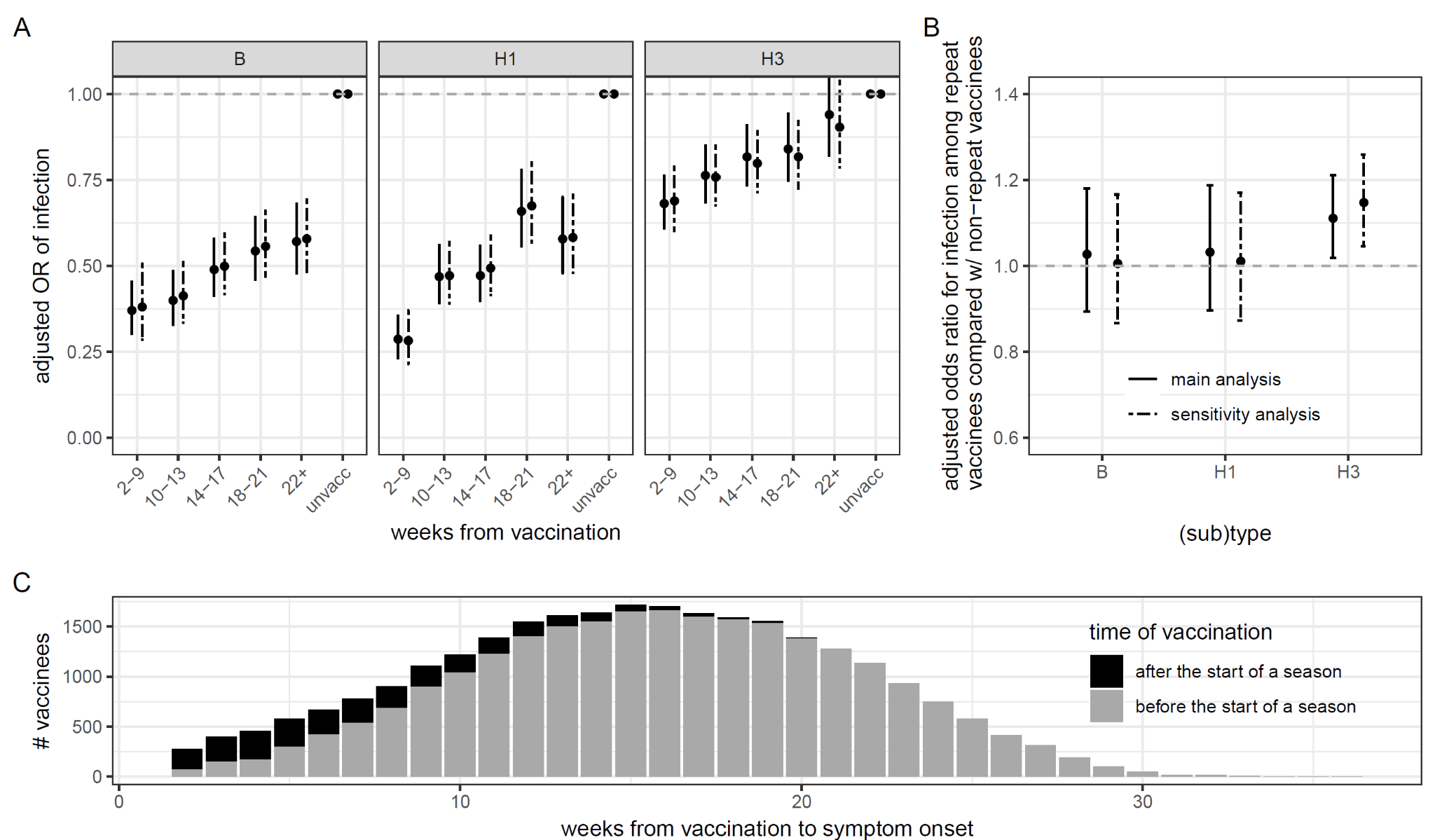

Figure 3.4: Impact of waning protection before (solid lines) and after (dashed lines) excluding individuals vaccinated after the start of a season (i.e., after the first symptomatic case was detected at each site). A) Estimated relative odds of Vaccine effectiveness estimates against clinical infection 2-9, 10-13, 14-17, and 22+ weeks after vaccination. B) Adjusted odds ratio for clinical infection comparing repeated vaccinees with non-repeated vaccinees. The solid lines show results that have been shown in Figure 1B in red. In the first two panels, error bars indicate 95% confidence intervals. C) Distribution of weeks from vaccination to symptom onset. Black represents individuals who were vaccinated after the start of a season and were thus excluded from this sensitivity analysis.

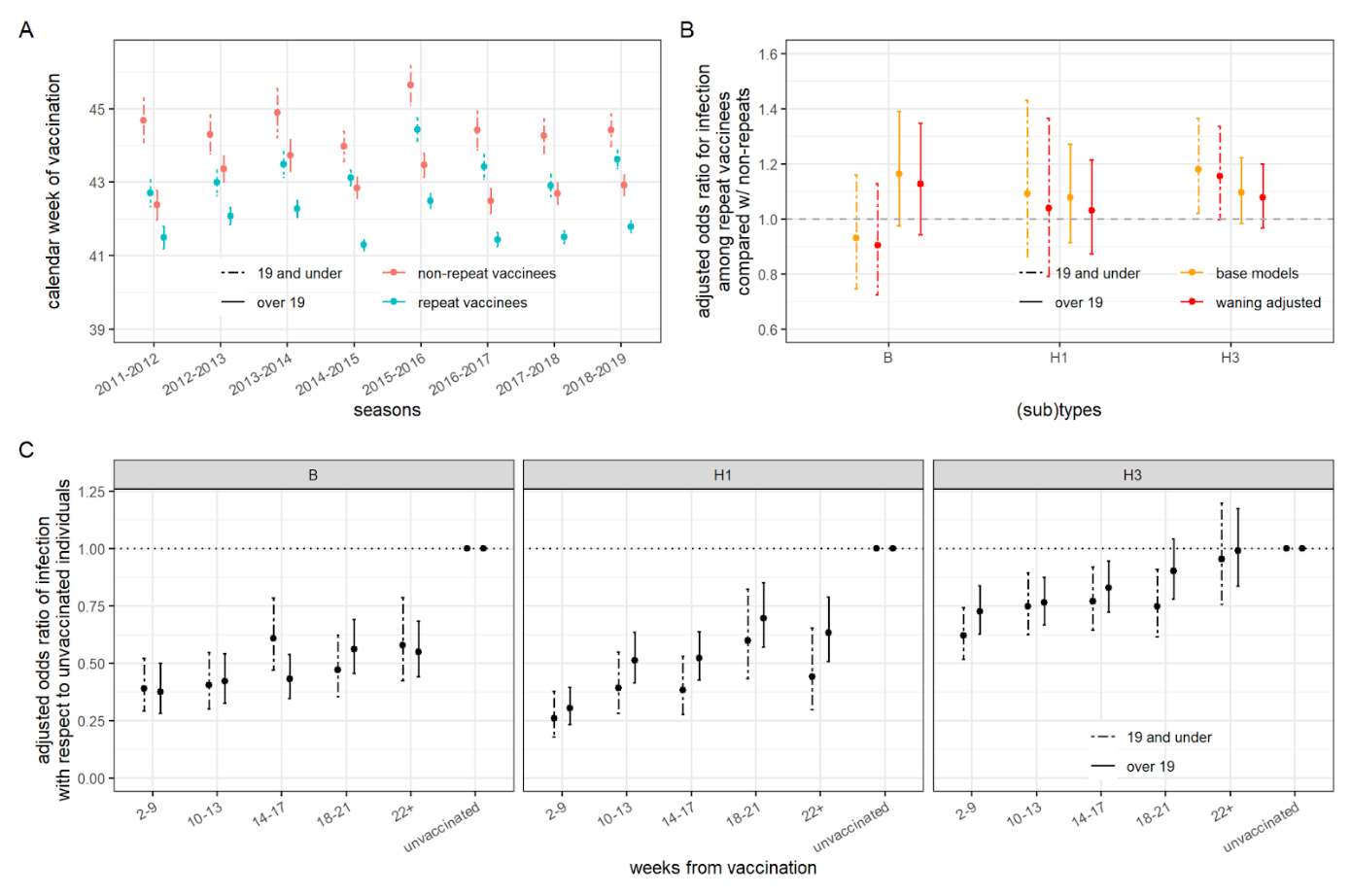

Figure 3.5. Impact of timing of vaccination and waning vaccine effectiveness on estimated effect of repeat vaccination, stratified by age group. A) Average calendar week of vaccination among repeat and non-repeat vaccinees over the study enrollment seasons. Repeat vaccinees consistently get vaccinated earlier than non-repeat vaccinees. B) Adjusted odds ratio for clinical infection among individuals vaccinated this season stratified based on whether the individuals were also vaccinated in the prior season (repeat vaccinees) or not (non-repeat vaccinees) before (yellow) and after (red) adjusting for the timing of vaccination within a season. Site- and season- specific data are shown in SFig 3.3). C) Adjusted odds ratio of clinical infection comparing individuals vaccinated 2-9, 10-13, 14-17, 18-21, and 22+ weeks before testing positive in the current season (but not in the previous season) with respect to those not vaccinated in either season.

Section 4. Application of inverse-probability weighted regression model

Because MCHS was the only site that had linked previous study enrollment and infection history for participants, only data from MCHS could be used to assess the impact of clinical infection history.

To determine how a clinical infection outcome in the current season is associated with clinical infection with the homologous (sub)types and separately with the heterologous (sub)types in any prior season, we assessed the odds ratio of clinical infection in the current season among 3 groups of individuals with reference to those whose last detected clinical infection was 1-2 seasons before the current season. The three groups are individuals whose last detected clinical infection was 3-5 seasons before the current season, 6-9 seasons before the current season, as well as individuals who had no documented infection during the enrollment seasons (i.e., since the 2007-2008 season) indicating that their most recent prior clinical infection was reported ≥ 9 seasons before the current season, dated back to seasons outside of our study period, or that they had no detected symptomatic infection. The reference group is individuals whose last detected clinical infection was 1-2 seasons before the current season. The other independent variables are sex, age category, comorbidity, calendar month of onset, influenza season, and current- and previous-season vaccination status. We excluded from this analysis individuals enrolled during the 2009-2010 season and the 2009 Pandemic season due to the atypical circulation of virus and vaccine types.

We then assessed whether clinical influenza virus infections in the previous season influenced the decision to vaccinate in the current season using logistic regression models. Only the vaccination status in the current season and the prior season were used because we wanted to use the same vaccination records that were collected by the other US Flu VE Network sites. The dependent variable is vaccination in the current season, and the independent variable is infection status of any (sub)type in the previous season. The model is stratified by vaccination status in the previous season. The model additionally adjusted for age group, sex, comorbidity, and an indicator of vaccination frequency (i.e., out of the three seasons immediately before the enrollment season, the number of seasons in which individuals were vaccinated). We excluded those who were vaccinated in the previous season after being clinically infected in that season. We assessed individuals’ tendency to switch vaccination status after clinical infections by age group, separately among individuals vaccinated and unvaccinated in the previous season.

Next, we estimated the effect of repeated vaccination after adjusting for the clinical infection status of any (sub)type in the previous season using IP-weighted logistic regression models. Clinical infection status in the previous season (*I_0_*) is both a confounder and a mediator of the joint effect of current- and previous-season vaccination status on infection in the current season (i.e., the joint effects of *V_0_* and *V_1_* on *I_1_* as illustrated in Supplemental Figure 2.1). To handle this treatment-confounder feedback, we used an inverse-probability weighted regression model [[52]](https://paperpile.com/c/kjBGeu/8YZ4), whereby adjustment for clinical infection status in the previous season is addressed by weighting while adjustment for baseline covariates *C* is made by regression.

Weights are calculated based on the inverse of each individual's probability of being vaccinated at each season, given their vaccination status in the previous season, infection status in the previous season, baseline covariates C (i.e., sex, age group, comorbidities, enrollment season) (i.e., the denominator of sw). These inverse-probability weights are then “stabilized” using the probabilities of being vaccinated given vaccination history and baseline covariates (i.e., the numerator of sw). Stabilized weights (vs. non-stabilized weights) generally resulted in narrower confidence intervals around the effect estimates. The numerator and denominator of the weights are estimated using logistic regression.

The general form of an individual’s stabilized weights *sw* in the current season can be expressed as

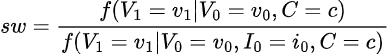

where $V_{0}$ and $I_{0}$ represent vaccination and infection status in the previous season, respectively, and $V_{1}$ represents vaccination in the current season. *C* is a vector of confounders (i.e., sex, age, comorbidities) measured at the underlying baseline of the two seasons being evaluated. Although comorbidities were only measured at the current season, we considered the presence of comorbidities a baseline covariate, assuming that it remained the same over the two seasons evaluated. *SZ* represents the influenza season at enrollment. For this analysis, infection status $I_{0}$=1 in the case of PCR-confirmed clinical infection of any influenza (sub)type, $V_{0}$=1 in the case of vaccination two weeks before symptom onset in the previous season, and $V_{1}$=1 in the case of vaccination at least two weeks before symptom onset in the current season.

Using IP-weighted logistic regression models, we then estimated relative odds of infection among repeat vaccinees compared with non-repeat vaccinees. The study outcome was (sub)type specific PCR-confirmed clinical infection in the current season. In the main analyses, the independent variables are an indicator for having been vaccinated 2-9, 10-13, 14-17, 18-21, or >21 weeks before symptom onset in this season, a dichotomous indicator for having been vaccinated only in the prior season, a dichotomous indicator for having been vaccinated in both the current and the prior season, age group, sex, influenza season, and comorbidities. 95%CI was generated using bootstrapping. Individuals who were not clinically infected in the previous season consisted of those who did not enroll in the previous season, those who were eligible but refused enrollment, and those who were enrolled but tested negative for influenza. We did not adjust for the calendar month of symptom onset in the model. Instead, to account for the possible influence of the timing and intensity of influenza epidemics each season on the timing of vaccination, we excluded individuals who were vaccinated before the start of a season (i.e., individuals who were vaccinated after the first 5% of PCR-confirmed cases presented symptoms in each season), under the assumption that the decision to vaccinate early in a season was not correlated with risk factors for infection (e.g., age and comorbidities). We accounted for waning vaccine protection in the main analyses, but we omitted it in sensitivity analyses for consistency with a typical longitudinal cohort analysis of a 2-season period, which would not condition on timing of vaccination as a post-baseline variable in the weighted outcome model. We excluded those who were eligible but refused enrollment in the previous season as a sensitivity analysis [[52]](https://paperpile.com/c/kjBGeu/8YZ4).

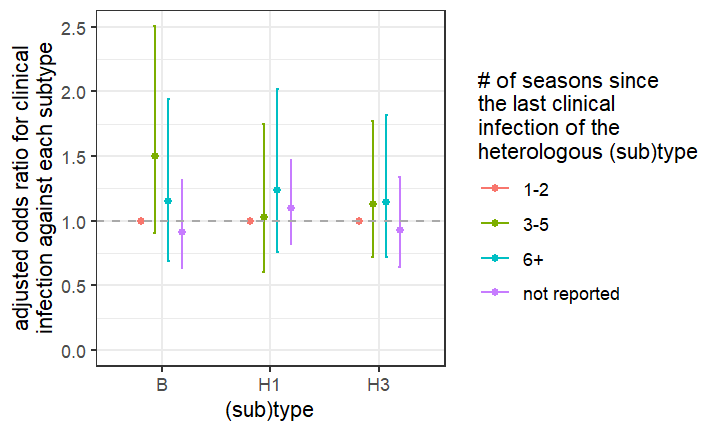

Figure 4.1: Protection conferred by infection of a heterologous subtype. Number of seasons from the last clinical infection of the heterologous subtype was not associated with odds of clinical infection. Error bars indicate 95% confidence intervals.

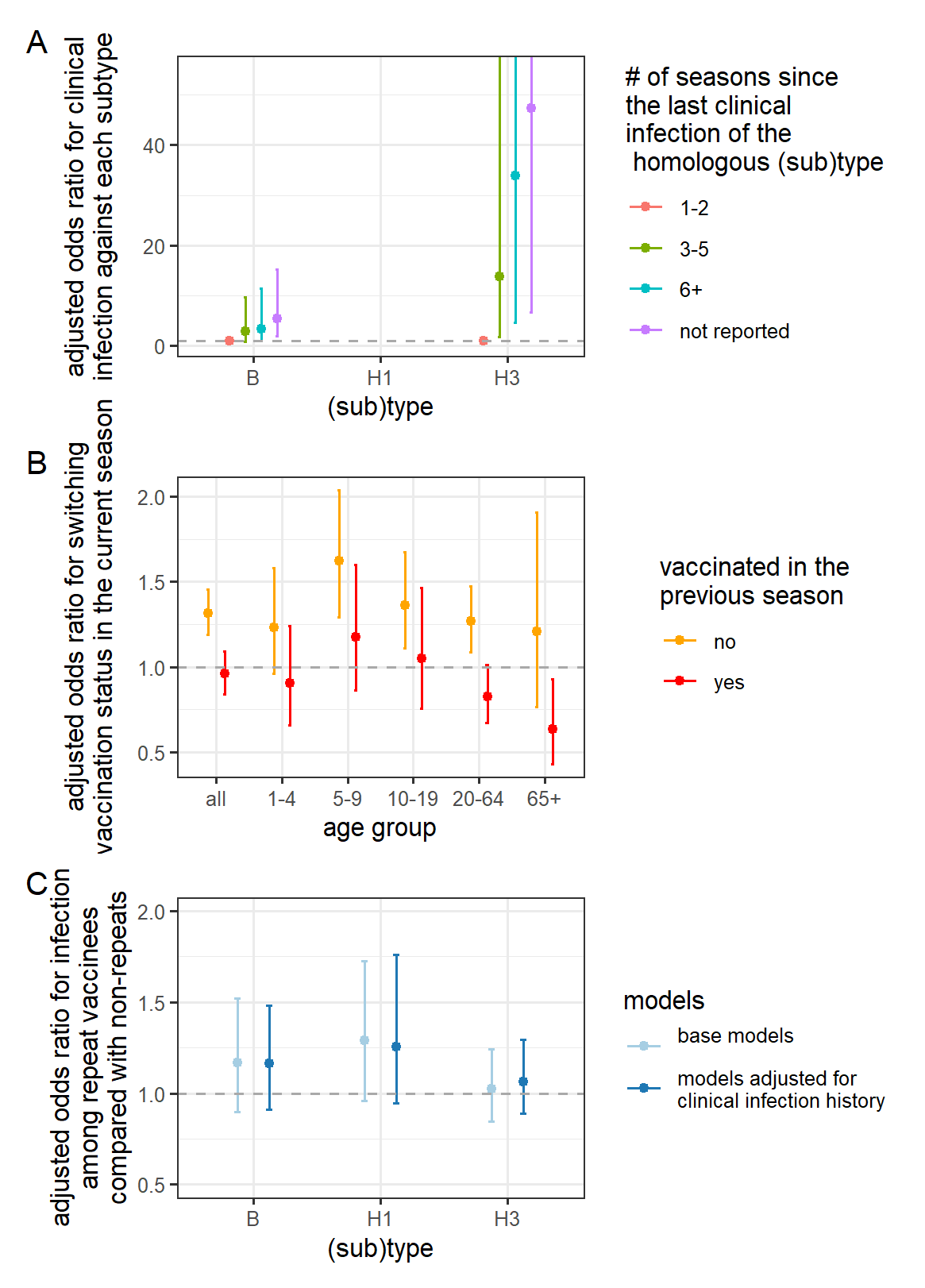

Figure 4.2: Impact of clinical infection on the estimated effect of repeated vaccination among participants from the Marshfield Clinic Health System excluding those who presented with acute respiratory illness but refused enrollment in the previous season. The results from this sensitivity analysis were qualitatively similar to the main analysis presented in Figure 2. A) Association between recent clinical infection and odds of current-season clinical infection. More distant clinical infections of the homologous subtype are associated with a higher odds of clinical infection. B) Tendency to switch vaccination status in the current season after infection in the prior season. Unvaccinated individuals were more likely to vaccinate in the current season if they were clinically infected in the previous season than if they were not. C) Estimated effect of repeated vaccination after adjusting for recent clinical infections. Adjusted odds ratio for clinical infection comparing repeated vaccinees with non-repeated vaccinees before (light blue) and after adjusting for the infection status in the previous season (dark blue) using inverse-probability weighting. Adjustment did not significantly impact the estimates. In all panels, error bars indicate 95% confidence intervals.

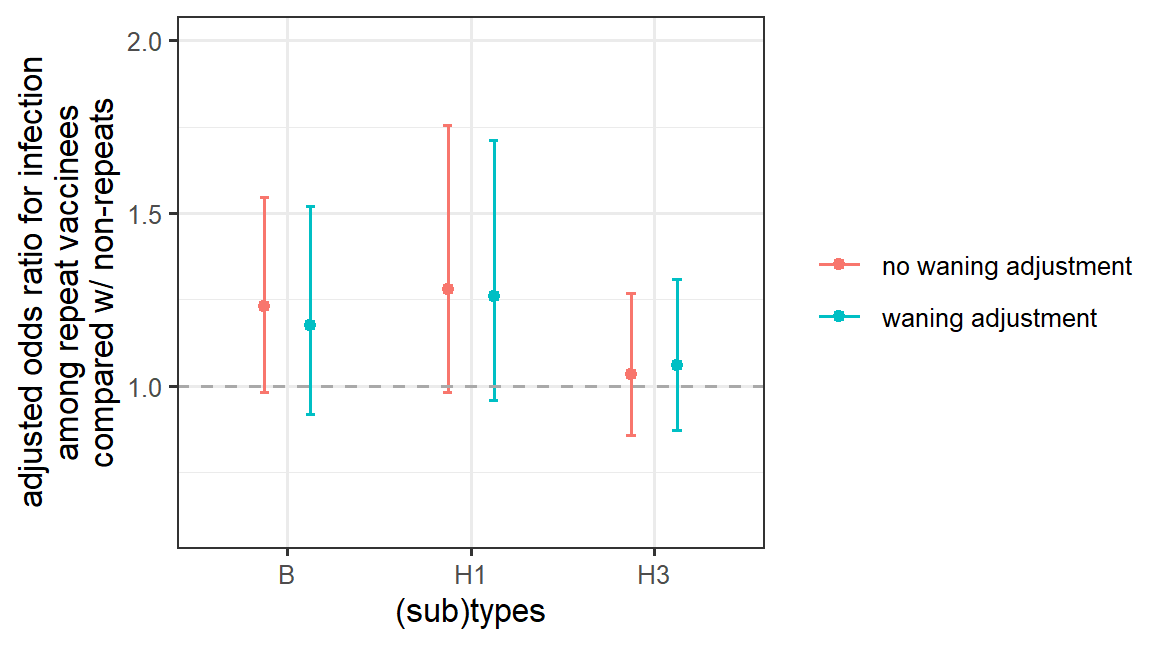

Figure 4.3. Adjusted odds ratio for clinical infection comparing repeat vaccinees with non-repeat vaccinees from the Marshfield Clinic Health System using inverse-probability weighting before (red) and after (blue) adjusting for the timing of vaccination within a season in the weighted outcome models.

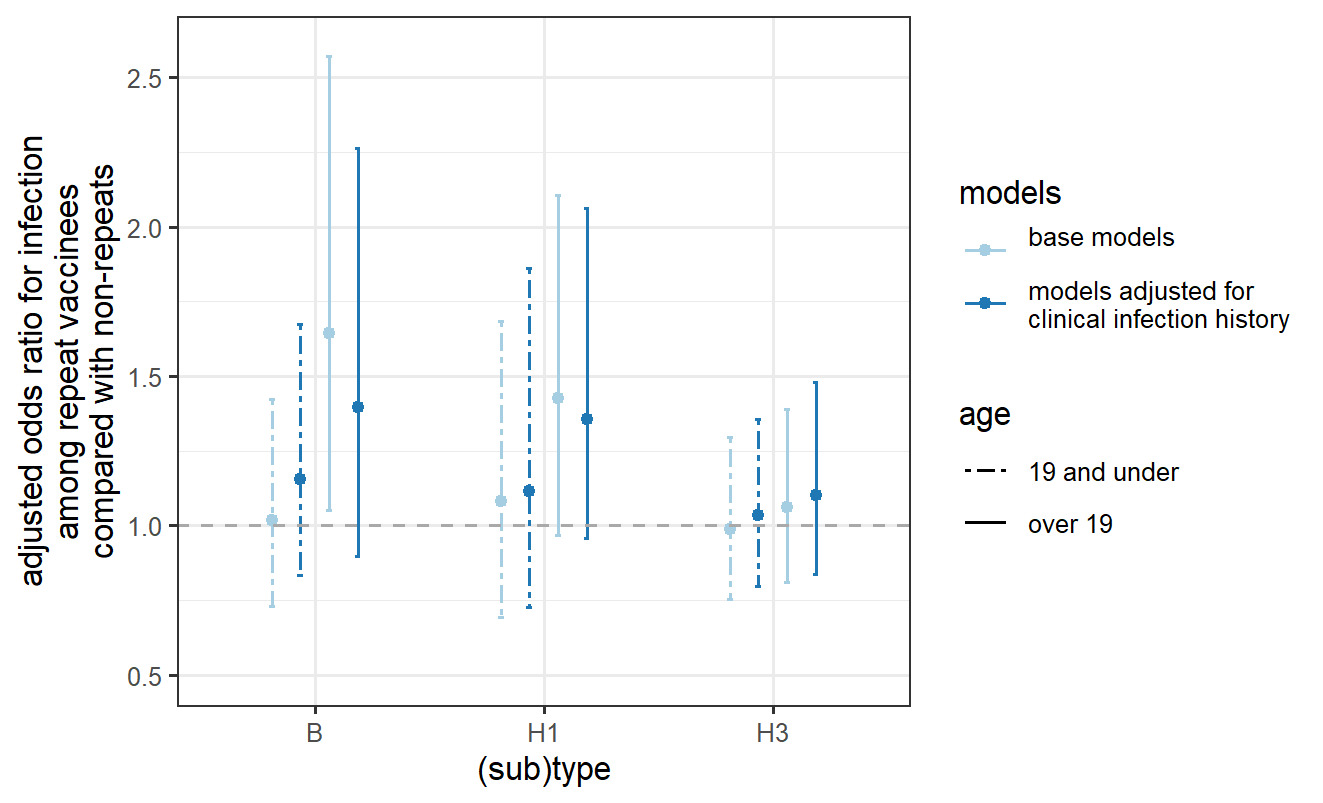

Figure 4.4. Adjusted odds ratio for clinical infection comparing repeat vaccinees with non-repeat vaccinees from the Marshfield Clinic Health System stratified by age group.

Section 5. Methods for calculating the extent to which subclinical and clinical infection contribute to the observed repeat vaccination effect

5.1 Detailed methods

To understand how subclinical infection history not detected by the US Flu VE Network may impact the estimated effect of repeated vaccination, we evaluated the proportion of repeat and non-repeat vaccinees who would have had to have been subclinically infected in the previous season to reproduce the observed estimates. To achieve this objective, we built a theoretical model and created a pseudo-population of repeat and non-repeat vaccinees with various infection statuses in the previous seasons. We then varied assumptions about the protection conferred by clinical and subclinical infection in the prior season against future infection in the various groups. That is, we assumed clinical infection in the previous season confers perfect protection against clinical infection in the current season, whereas subclinical infection in the previous season confers partial protection in the current season. We also assumed vaccination status in the previous season does not affect the odds of infection in the current season. Based on estimates from studies in the US [[53]](https://paperpile.com/c/kjBGeu/UP8I), we assumed a 1% clinical attack rate among vaccinated individuals (denoted by *a*) and 2% among unvaccinated individuals (denoted by *x*). We also assumed a 1.5% current-season clinical attack rate among vaccinees not infected in the previous season (denoted by *γθ*). Detailed methods and assumptions are shown below. We illustrated how the results are consistent with both the infection block hypothesis and the hypothesis of enhanced vaccine immunogenicity post-infection under different interpretations of the same parameters.

Table 5.1: The table illustrates how the rates of subclinical and clinical infection can drive the estimated effect of repeated vaccination through the infection block hypothesis and the hypothesis of enhanced immunogenicity post-infection

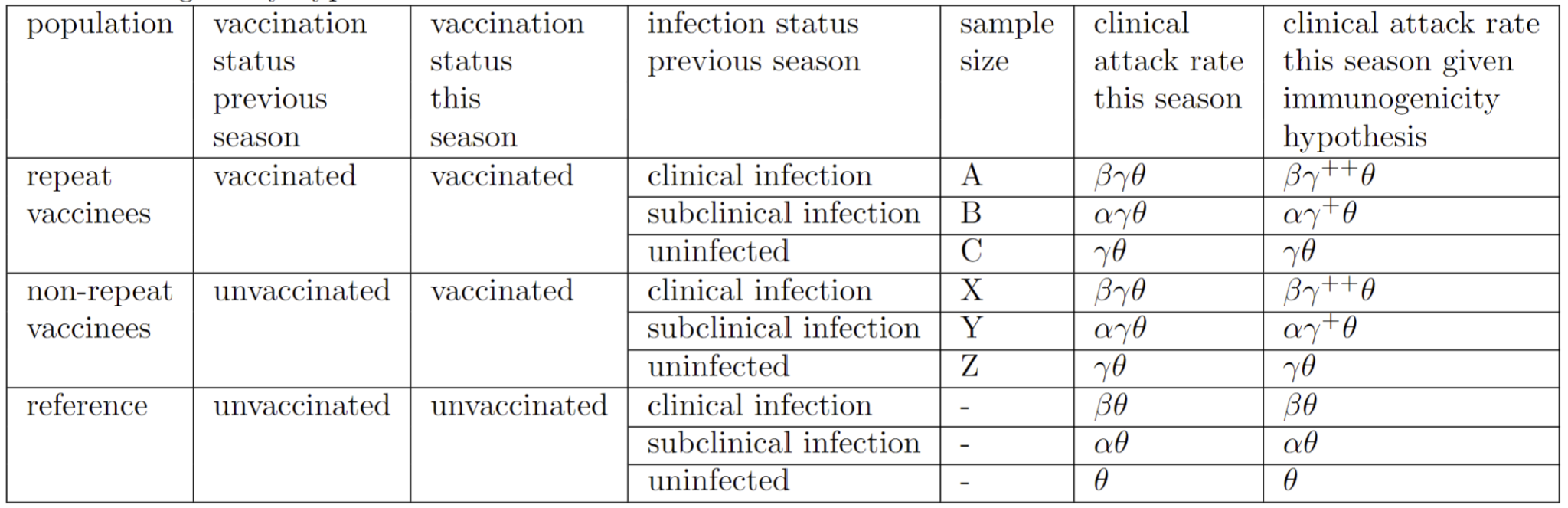

Table 5.1 shows the attack rate and population size of repeat and non-repeat vaccinees given varying exposure and infection status in the previous seasons.

Let $\theta$ be the current-season clinical attack rate among individuals who were neither vaccinated nor infected (clinically or subclinically) in the prior season, 1-α be the effectiveness of subclinical infections that occurred in the previous season against clinical infection in the current season, 1-β be the effectiveness of clinical infections that occurred in the previous season against clinical infection in the current season, and 1-γ be the current-season VE against clinical infection. If we assume that vaccines were more effective among individuals who were infected in the previous season than those who were not infected, current-season VE against clinical infection would increase to 1-γ^++^ and 1-γ^+^ among those clinically and subclinically infected in the previous season, respectively (0 <*γ*^++^<*γ*^+^<*γ*<1).

We assumed that the effectiveness of subclinical or clinical infection that occurred in the previous season in preventing infection in the current season was not influenced by previous-season vaccination status. For example, repeat vaccinees’ current-season attack rates among individuals who had clinical infection in the previous season are the same as that among non-repeat vaccinees. In addition, those not infected in the previous season may have escaped infection for various reasons: they were not exposed, or they were exposed but protected by natural or vaccine-induced protection. We assumed the clinical attack rate to be the same among those who were not infected in the previous season regardless of their exposure history prior to the previous season.

Table 5.2. Calculation of the size of repeat and non-repeat vaccinees who are infected and uninfected in the current season. AR represents attack rate.

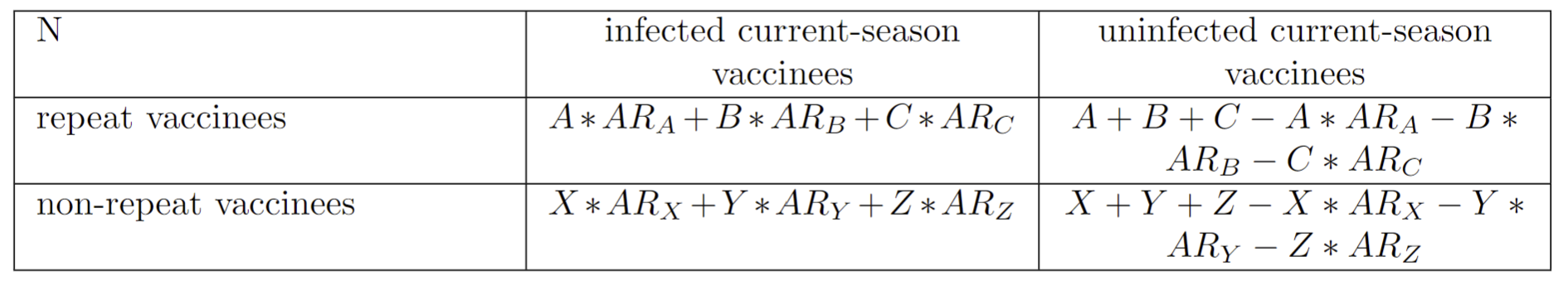

The odds ratio for clinical infection comparing repeat vaccinees with non-repeat vaccinees is:

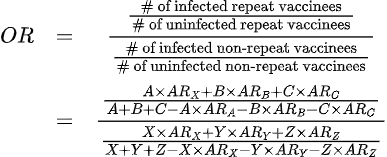

Let $a=\frac{A}{A+B+C}, b=\frac{B}{A+B+C},x=\frac{X}{X+Y+Z}, y=\frac{Y}{X+Y+Z}$ . We assumed recent clinical infections conferred perfect protection against clinical infections in the current season (i.e., *AR*_A_ = 0, *AR*_X_ = 0). Then the OR can be simplified to:

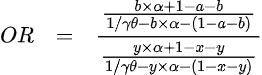

which can be rearranged to obtain a relationship between the fraction of repeat vaccinees who were subclinically infected in the previous season (denoted by y) and the fraction of non-repeat vaccinees who were subclinically infected in the previous season (denoted by b):

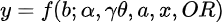

For the main analyses, we illustrated this relationship in Figure 3A assuming a 1% clinical attack rate among vaccinated individuals (*a*) and 2% among unvaccinated individuals (*x*), both are weighted averages by their infection status in the previous season. As a result, the current-season clinical attack rate among vaccinees infected in the previous season, although not specified, should be lower than 1% and can be calculated based on our assumption of a 1.5% current-season clinical attack rate among vaccinees not infected in the previous season (*γθ*). We illustrated this relationship under 3 levels of protection after subclinical infection (i.e., *α* = 30%, 50%, and 70%).

Table 5.3. Descriptions of parameters and estimates used in the models

| Notation | Description | Parameter values in the main analyses | Parameter values in the sensitivity analyses for high-incidence settings | Parameter values in the analyses where we tested the impact of enhanced vaccine immunogenicity post-infection |
| --- | --- | --- | --- | --- |
| a | Clinical attack rate among vaccinees | 1% | 3% | 1% |
| b | Subclinical attack rate among current season vaccinees who were vaccinated in the prior season as shown on the x-axis of Figure 3A in the main text | Explored the range between 0 and 50% | Explored the range between 0 and 50% | Explored the range between 0 and 50% |
| x | Clinical attack rate among unvaccinated individuals | 2% | 6% | 2% |
| y | Subclinical attack rate among current-season vaccinees who were unvaccinated in the prior season as shown on the y-axis of Figure 3A in the main text | Explored the range between 0 and 50% | Explored the range between 0 and 50% | Explored the range between 0 and 50% |
| *1-α* | Level of protection after subclinical infection | 30%, 50%, 70% | 30%, 50%, 70% | 30%, 50%, 70% |
| *γθ* | Current-season clinical attack rate among vaccinees not infected in the previous season. (This rate is higher than the clinical attack rate for vaccinees overall, which is represented by *a*, because it includes only vaccinees without additional protection from recent infection.) | 1.5% | 5% | 1.5% |
| *1-γ* | Overall vaccine effectiveness against clinical infection | 50% | 50% | 50% |
| *1-β* | The effectiveness of clinical infections in the prior season against clinical infection in the current season | Not relevant | Not relevant | Not relevant |
| *θ* | Clinical attack rate among individuals not vaccinated either in the current or the prior season and was uninfected in the prior season | 3% | 10% | 3% |
| *OR* | Odds ratio of infection comparing repeat and non-repeat vaccinees | 0.7, 0.8, 0.9, 1, 1.1, 1.2, 1.3 | 0.7, 0.8, 0.9, 1, 1.1, 1.2, 1.3 | 1.1 |
| 1 *− γ*^+^ | Enhanced overall vaccine effectiveness against clinical infection assuming that recent infection boosts vaccine immunogenicity and thus vaccine-induced effectiveness | Not relevant | Not relevant | Explored the range between 50% to 100% |

5.2 Additional results

Previously we found that in the US Flu VE Network, the odds of infection against A/H3N2 (and type B) among repeat vaccinees compared with non-repeat vaccinees is about 1.1. When OR is 1.1, we found that the rate of subclinical infection among non-repeat vaccinees in the previous season needs to be consistently larger than the rate among repeat vaccinees under a range of reasonable parameter estimates (i.e., fraction of all repeat vaccinees who were subclinically infected were under 30%) to match this OR.

In Figure 3A of the main analyses, we show that for example, if the protection against future clinical infections after a subclinical infection is 30% (the first panel) and 20% of repeat vaccinees were subclinically infected in the prior season (the x-axis), then 45% of non-repeat vaccinees would have to have been subclinically infected in the prior season (the y-axis) to observe the estimated effect of repeated vaccination in the US Flu VE Network (i.e., odds ratio for clinical infection comparing repeat vaccinees with non-repeat vaccinees against A/H3N2 or type B of 1.1).

 Larger differences in subclinical attack rates between the two groups or stronger protection against clinical infection after subclinical infection would each lead to a greater excess of clinical infections in the current season among repeat vaccinees compared with non-repeat vaccinees (Figure 3A), leading to increased odds of clinical infection among repeat vaccinees, consistent with the infection block hypothesis.

When repeat and non-repeat vaccinees experienced subclinical infections in the previous season at the same rate, a scenario represented by the diagonal lines in Figure 3A, the higher rate of clinical infection among non-repeat vaccinees compared with repeat vaccinees (1% vs. 2% based on our assumption) could elevate OR to only about 1.01 (vs. the observed ORs of about 1.1 in the CDC Flu VE Network).

Although we did not measure the rate of subclinical infections through serological studies, we found in theoretical analyses that relative protection would be reduced in repeat vaccinees to the extent estimated in the US Flu VE Network if they experience a lower rate of partially protective subclinical infections in the prior season compared to non-repeat vaccinees. We observed that when the protection against future clinical infection after subclinical infections is 70%, the rate of subclinical infections would have to be lower in repeat vaccines than in non-repeat vaccines by about 10 percentage points to create the observed increase in clinical infection risk among repeat vaccinees. This absolute difference would increase to about 25 percentage points if the protection against future clinical infection is approximately 30%. This finding provides an explanation for increased risk after repeated vaccination after adjusting for clinical infection history alone. We demonstrated mathematically that smaller differences in subclinical infection rates could generate observed differences in VE if subclinical infection also substantially enhances the immunogenicity of vaccination in the next season.

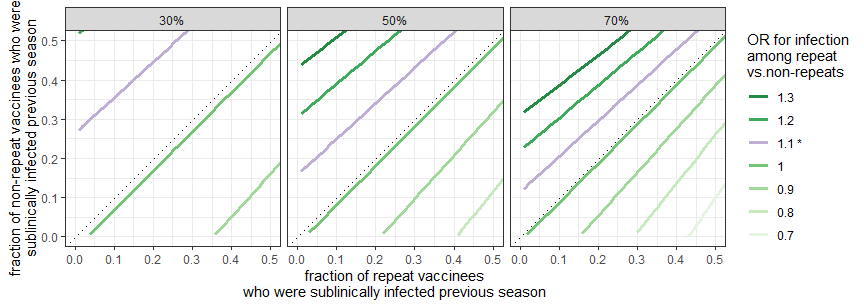

Figure 5.1: Subclinical infection might be able to explain the effect of repeated vaccination. The figure shows the fraction of repeat and non-repeat vaccinees who would need to have been subclinically infected in the previous season to reproduce the estimated effect of repeated vaccination in the US Flu VE Network. See Supplementary Section 5 for detailed methods. The estimated effect of repeated vaccination (OR=1.1) in the US Flu VE network is colored in purple. The results shown here are generated assuming vaccine effectiveness against clinical infection is 50%; clinical attack rate among vaccinees in a season is 1%; current-season clinical attack rate among the subset of current-season vaccinees not infected in the previous season is 1.5%; and clinical infection in the previous season perfectly protects against clinical infection in the following season. Each facet represents a predetermined protection against clinical infection after subclinical infection (i.e., 30%, 50%, 70%). The legend represents the estimated effect of repeated vaccination given the difference in subclinical attack rate among repeat vaccinees (x-axis) and non-repeat vaccinees (y-axis) and assumptions stated above. Only the plausible range of subclinical attack rate among repeat vaccinees (x-axis) and non-repeat vaccinees (y-axis; 0-50%) are shown in the figure.

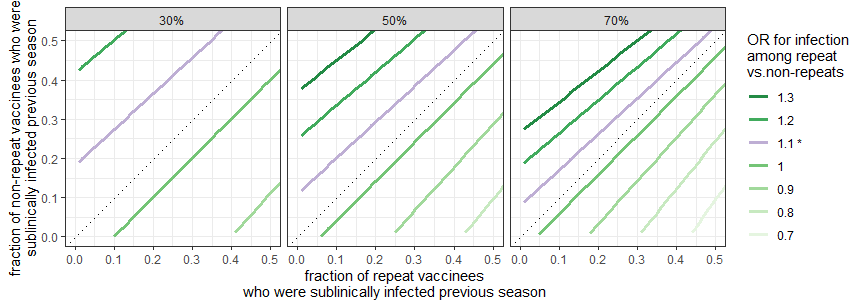

Figure 5.2. Sensitivity analysis on the results shown in Figure 5.1 using parameters that resemble values from high incidence settings. We assumed the clinical attack rate among vaccinees in a season is 3%, and the current-season clinical attack rate among the subset of current-season vaccinees not infected in the previous season is 5%. All other assumptions remain the same as the analyses shown in Figure 5.1

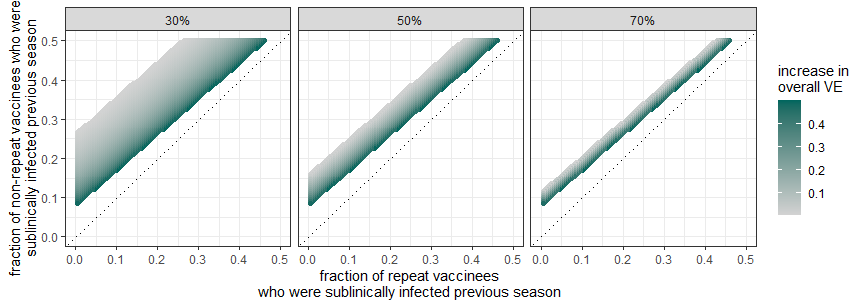

Figure 5.3: The absolute increase in VE from a baseline VE (shown in the legend) of 50% needed to reproduce the observed estimated effect of repeated vaccination in the US Flu VE Network (OR=1.1). The estimated increase in VE against clinical infection varies by the effectiveness against future clinical infections after subclinical infection (i.e., 30%, 50%, 70%, one in each facet). The uncolored portion of the figure represents the population where a boost in VE after infection will not generate the observed effect of repeated vaccination (OR of 1.1). The results shown here are generated assuming the current-season clinical attack rate among vaccinees in a season is 1%, and the current-season clinical attack rate among the subset of current-season vaccinees not infected in the previous season is 1.5%. We also assumed that clinical infection in the previous season perfectly protects against clinical infection in the current season.

Section 6. Exploring the mediation effect of prior-season clinical infection

We adapted the model described in section 5 of the supplementary material to explore the possibility that prior-season clinical infection acts as an important mediator between prior-season vaccination and risk of current-season clinical infection.

Unlike the previous section, we assumed that prior clinical infection does not confer perfect protection against future clinical infection (i.e., *1-β > 0*). We kept other assumptions from the previous section: We assumed a 1% clinical attack rate among vaccinated individuals (a) and 2% among unvaccinated individuals (x), both are weighted averages by their infection status in the previous season. As a result, the current-season clinical attack rate among vaccinees *infected* in the previous season, although not specified, should be lower than 1% and can be calculated based on our assumption of a 1.5% current-season clinical attack rate among vaccinees *not infected* in the previous season (*γθ).* We illustrated this relationship under 3 levels of protection after subclinical infection (i.e., *α* = 30%, 50%, and 70%) and combinations of a range of subclinical attack rate among current-season vaccinees who were vaccinated (b) and unvaccinated (y) in the prior season.

In short, under a range of parameters and assumptions, the protection conferred by clinical infection in the prior season led to little variation in the estimated effect of repeated vaccination in the theoretical model, suggesting that prior-season clinical infection is unlikely to be an important mediator in this relationship. Regardless of the protection after subclinical infection (*α*), prior-season subclinical attack rate among repeat and non-repeat vaccinees (b and y), estimated ORs indicating the effect of repeated vaccination (y-axis in the following figure) vary little with the effectiveness of prior-season clinical infection (x-axis in the following figure).

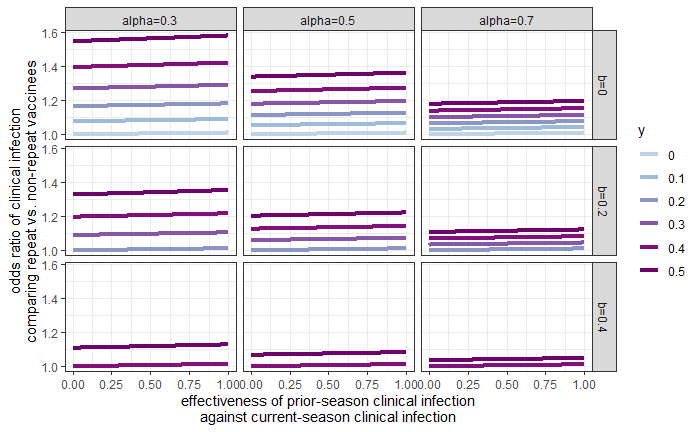

Figure 6.1: Potential of prior-season clinical infection for mediating the effect of prior-season vaccination and the odds of current-season clinical infection. 1-*α* represents the level of protection against clinical infection conferred by prior-season subclinical infection. b and y represent the subclinical attack rate among current-season vaccinees who were vaccinated (b) and unvaccinated (y) in the prior season, respectively. The effectiveness of prior-season clinical infection against current-season clinical infection as shown on the x-axis is denoted by *1-β*. Lack of variation in the odds ratio indicating the estimated effect of repeated vaccination (shown on the y axis) indicates that prior-season clinical infection is unlikely to be an important mediator of the effect of prior-season vaccination on odds of current-season clinical infection.

Additional Reference

.	51.	Li KQ, Shi X, Miao W, Tchetgen ET. Double Negative Control Inference in Test-Negative Design Studies of Vaccine Effectiveness. ArXiv **2022**; Available at: <https://www.ncbi.nlm.nih.gov/pubmed/35350548>.

52.	[Robins JM, Hernán MÁ, Brumback B. Marginal Structural Models and Causal Inference in Epidemiology. Epidemiology. 2000; 11:550–560. Available at:](about:blank) <http://dx.doi.org/10.1097/00001648-200009000-00011>.

53.	Jackson ML, Jackson LA, Kieke B, et al. Incidence of medically attended influenza infection and cases averted by vaccination, 2011/2012 and 2012/2013 influenza seasons. Vaccine. 2015; 33:5181–5187. Available at: http://dx.doi.org/10.1016/j.vaccine.2015.07.098.
